# Genomic surveillance of *Salmonella enterica* serotype Minnesota strains from poultry products imported into South Africa

**DOI:** 10.1101/2025.04.16.25325939

**Authors:** Vishnu Raghuram, Thendo Mafuna, Vignesh Ramnath, Hadrien Gourlé, Josefin Blom, Kudakwashe Magwedere, Laura M. Carroll, Itumeleng Matle

## Abstract

*Salmonella enterica* serotype Minnesota (*S.* Minnesota) has recently emerged as a predominant serotype in poultry farming operations. Genomic surveillance efforts concentrated primarily in Europe have been used to evaluate food safety risks associated with *S.* Minnesota in imported poultry/poultry products. However, the burden imposed by *S.* Minnesota on consumers in sub-Saharan Africa is not understood. Here, we used whole-genome sequencing (WGS) to characterize 36 *S.* Minnesota strains from raw poultry imported into South Africa, specifically: (i) 11 strains isolated at port-of-entry, and (ii) 25 strains from imported poultry in South African supermarkets. While all 36 strains belonged to the same sequence type (ST548), multiple ST548 lineages were present among poultry products. Comparison of the 36 strains sequenced here to all publicly available, high-quality ST548 genomes (*n* = 228, from Enterobase) identified several closely related public genomes (<30 core SNPs), including strains isolated previously from South American poultry imported into the United Kingdom. Notably, a cluster consisting of 14 highly similar genomes sequenced here (0 core SNPs) uniquely possessed *bla*_CTX-M-8_. A search of plasmids in public databases, alongside antimicrobial resistance (AMR) genes from >1.9 million bacterial genomes, revealed that this cluster harbored *bla*_CTX-M-8_ on an IncI1 plasmid, which we hypothesize was acquired recently, from *Escherichia coli*. Overall, our study provides insight into the intercontinental dissemination of *S.* Minnesota and its associated AMR determinants via the global poultry trade.

**Impact Statement:** Raw poultry exports have disseminated *S.* Minnesota internationally, and several countries (primarily in Europe) have used WGS to characterize *S.* Minnesota strains from imported poultry. While many African countries also import significant quantities of poultry, very little is known about *S.* Minnesota in South Africa, let alone sub-Saharan Africa. Here, we detect multiple *S.* Minnesota ST548 lineages at two control points in South Africa (port-of-entry and supermarkets), including emerging AMR lineages. Notably, we found that all *S.* Minnesota genomes from South Africa were closely related to poultry/food-associated *S.* Minnesota genomes from the United Kingdom (UK) or South America. Recent WGS-based studies from the UK have posited that *S.* Minnesota from poultry is unlikely to cause illness, should it reach UK consumers. While no links to clinical cases in South Africa were observed here, this could be due to data gaps, as the vast majority of *Salmonella* WGS efforts are concentrated in Europe and North America. Our study highlights the important role that genomic surveillance plays in mitigating food safety risks associated with the global agro-food trade and showcases the importance of local pathogen surveillance initiatives.

**Data Summary:** WGS data generated in this study is available under NCBI BioProject accession PRJNA1230142. NCBI BioSample and Sequence Read Archive (SRA) accessions for newly sequenced genomes, as well as for the publicly available genomes used in this study, are available in the Supplementary Material. All code for running software is described in the article, and additional data analysis code is available via GitHub (https://github.com/VishnuRaghuram94/SEPI). Supplementary Tables S1-S3 are available alongside the article. All larger supplementary datasets (Dataset S1-S17) and intermediate files are available via doi: 10.5281/zenodo.15063662.

## INTRODUCTION

Foodborne zoonotic pathogen *Salmonella enterica* imposes a massive burden on global public health [1, 2]. Each year, *Salmonella* is estimated to cause 93.8 million cases of gastroenteritis globally, the vast majority of which (>85%) are attributed to the consumption of contaminated food [3]. Contaminated poultry in particular is widely regarded to be a major source of *Salmonella* illnesses [1, 4–6]. In the United States (US), for example, chicken and turkey products have been estimated to account for nearly a quarter of foodborne *Salmonella* illness cases [7], with meat and poultry products serving as the most common sources of fatal foodborne illness cases (caused primarily by *Listeria* or *Salmonella*) [8, 9].

While poultry can serve as a reservoir for a range of *Salmonella* serotypes [10–12], *Salmonella enterica* subspecies *enterica* serotype Minnesota (*S.* Minnesota) has recently emerged as a serotype of concern in Brazilian poultry farming operations [4, 13, 14]. First reportedly isolated from a turkey in Minnesota (US) in 1936 [15], *S.* Minnesota is a relatively rare serotype globally [4, 13]. However, in Brazil, *S.* Minnesota has recently emerged as a predominant serotype in poultry [4, 14, 16, 17]. It has been hypothesized that a combination of factors contributed to *S.* Minnesota’s rapid rise in Brazil, including the 2003 introduction of a vaccine against *S.* Enteritidis (the predominant serotype in Brazilian poultry at the time), antimicrobial use, and antimicrobial resistance (AMR) [4, 13, 18, 19].

From a genomic surveillance perspective, a great deal of effort has gone into monitoring the spread of S. Minnesota and its associated AMR determinants in Brazil [4, 14, 17, 20].

However, due to Brazil’s status as a leading global poultry exporter, S. Minnesota can be disseminated to other countries via imported Brazilian poultry/poultry products [4, 13]. As such, several other countries have employed whole-genome sequencing (WGS) to characterize S. Minnesota strains from Brazilian poultry imports [4, 13, 21, 22]. Notably, these efforts have been largely confined to Europe (e.g., the United Kingdom [UK], Portugal) [4, 21], and only recently, Saudi Arabia [13]. Very little is known about S. Minnesota in sub-Saharan Africa, and WGS-based studies of S. Minnesota from imported poultry have not been conducted in Africa, despite the fact that many African countries also import significant quantities of Brazilian poultry/poultry products [23].

Port-of-entry pathogen surveillance efforts in South Africa have highlighted the potential food safety risks associated with imported poultry products [24]. Given that (i) S. Minnesota has become relatively prevalent in imported poultry products tested at South African port-of-entry [24], and (ii) Brazil has been the largest exporter of poultry meat to South Africa since 2001 [25], it is apparent that poultry products imported from Brazil may serve as a vehicle by which S. Minnesota can enter South Africa’s domestic food system. However, the extent to which this has occurred has not been studied. Here, we used WGS to characterize 36 S. Minnesota strains isolated from raw poultry imported into South Africa from Brazil. By comparing our novel genomes to hundreds of relevant high- quality, publicly available S. Minnesota genomes from around the world, we provide insight into the dissemination of S. Minnesota and its associated AMR determinants in South Africa.

## METHODS

### Study design and source of isolates

This retrospective study was conducted using laboratory-confirmed *Salmonella* isolates stored at the Agricultural Research Council – Onderstepoort Veterinary Research (ARC- OVR) General Bacteriology Laboratory. The isolates were recovered from culture samples obtained during routine-controlled inspections at ports of entry (POEs), routine diagnostic services, and research studies at supermarkets. Sampling was not part of this study; however, isolates were sent to the ARC-OVR General Bacteriology Laboratory following the protocol recommended in Veterinary Procedural Notification (VPN) 56, accompanied by all necessary documentation and metadata. This included details such as the source of isolation, country or location of origin, and date of isolation or sample collection, among other relevant information.

### Sample isolation, DNA extraction, and WGS

Bacterial isolation and identification were carried out following the guidelines outlined in “Microbiology of the Food Chain: Horizontal Method for the Detection, Enumeration, and Serotyping of *Salmonella*” (ISO 6579-1:2017) [26]. Briefly, genomic DNA was extracted from overnight cultures using the High Pure PCR Template Preparation Kit (Roche, Germany) in accordance with the manufacturer’s instructions. WGS of the isolates was conducted at the Biotechnology Platform of the Agricultural Research Council (ARC), Onderstepoort, South Africa. DNA libraries were prepared using TruSeq DNA Library Preparation Kits (Illumina, San Diego, CA, USA) and sequenced on the HiSeq 2500 platform (Illumina, San Diego, CA, USA), according to the manufacturer’s instructions.

### Quality control, pre-processing, and assembly of WGS data

Trimming/filtering of raw Illumina paired-end reads was performed using fastp v0.23.4 with default parameters [27]. The resulting trimmed reads were assembled using Shovill v1.1.0 with the ‘--assembler skesà parameter (otherwise default parameters; https://github.com/tseemann/shovill) [28]. Quality control of the resulting assemblies was performed using: (i) QUAST v5.3.0 [29], with the complete *Salmonella enterica* sequence type 548 (ST548) str. ATCC 49284 genome as the reference (‘-r’ option, NCBI Assembly accession GCA_000486855.2; see section “*In silico* serotyping and sequence typing” below for sequence typing details) [30]; (ii) CheckM v1.2.2 [31], using the ‘lineage_wf’ workflow with default parameters (**Dataset S1**).

### *In silico* serotyping and sequence typing

*In silico* serotyping of assembled genomes (see section “Quality control, pre-processing, and assembly of WGS data” above) was performed using (i) SISTR v1.1.1 (default parameters) [32] and (ii) SeqSero2 v1.3.1 (‘-m k -t 1’; otherwise default parameters) [33]. *In silico* seven-gene multi-locus sequence typing (MLST) was performed using mlst v2.9 (https://github.com/tseemann/mlst) with default parameters (**Dataset S1**), assigning all genomes sequenced here to ST548.

### Acquisition of publicly available WGS data and metadata

All publicly available ST548 genomes (see section “*In silico* serotyping and sequence typing” above) and associated metadata were retrieved from Enterobase (*n* = 595 ST548 genomes, accessed 26 October 2022) [34, 35]. Samples were filtered to include only those associated with a BioProject [36] and for which a NCBI Assembly accession and year of isolation were available. In addition, only assemblies with (i) N50 > 50 Kbp, (ii) number of contigs (>=200bp) < 200, (iii) SISTR1 serotype = SeqSero2 serotype = “Minnesota”, and (iv) species percentage >= 85% were retained (assembly statistics obtained via Enterobase). This search resulted in a total of 229 publicly available genomes, which were downloaded from NCBI’s Assembly database (accessed 18 July 2024; **Dataset S2**).

To conduct a rough assessment of overall genome similarity among the 229 publicly available ST548 genomes, a Mash distance-based tree [37] was constructed using mashtree v1.4.6 [38], using ‘--genome-size 4716019’ (i.e., the average length of the 229 publicly available genomes). Inspection of the resulting Mash distance-based tree revealed a single outlier genome (NCBI Assembly accession GCA_011665355.1); this outlier genome was relatively distant from all other publicly available ST548 genomes and was thus not included in further analyses (**Fig S1**). The remaining 228 publicly available ST548 genomes were combined with the 36 newly sequenced genomes (see section “Sample isolation, DNA extraction, and WGS” above) to form the full dataset used in subsequent analyses (*n* = 264 total ST548 genomes, referred to hereafter as the “full ST548 dataset”; **Table S1**).

### AMR determinant and plasmid replicon detection

All 264 genomes in the full ST548 dataset (see section “Acquisition of publicly available WGS data and metadata” above) were screened for AMR determinants using AMRFinderPlus v3.12.8 [39, 40], with the ‘-O Salmonella --plus’ options and database version 2024-05-02.2 (otherwise default parameters; **Dataset S3**). ABRicate v1.0.1 (https://github.com/tseemann/abricate) was used with the PlasmidFinder database (‘--db plasmidfinder’, otherwise default parameters) to identify plasmid replicons in each genome (PlasmidFinder version 2023-Nov-4; **Dataset S4**) [41].

### Pan-genome analysis

All 264 genomes in the full ST548 dataset (see section “Acquisition of publicly available WGS data and metadata” above) were annotated using Bakta v1.8.2 with database version 5.0 [42]. The resulting .gff3 files were used as input for the pan-genome estimation software Panaroo v1.3.4 [43] with the following parameters: ‘-f 0.5 --core_threshold 0.95 --remove-invalid-genes --clean-mode strict -a pan‘. The resulting ‘gene_presence_absence.Rtab’ file was imported into R v4.4.0 (https://www.R-project.org/) and filtered to contain only intermediate genes (i.e., genes present in <95% and >5% of the population; **Dataset S5**). The intermediate gene presence/absence matrix was supplied as input to the R package umap v0.2.10.0 (https://CRAN.R-project.org/package=umap), and the resulting Uniform Manifold Approximation and Projection for Dimension Reduction (UMAP) [44] was visualized using ggplot2 v3.5.1 [45].

### Core SNP calling and phylogeny construction

Pairwise average nucleotide identity (ANI) values were calculated between all 264 genomes in the full ST548 dataset (see section “Acquisition of publicly available WGS data and metadata” above) using skani v0.2.0 [46] with the ‘--min-af 0’ option (otherwise default parameters; **Dataset S6**). The genome with the highest average ANI relative to all other genomes was selected as the reference genome for reference-based SNP calling (NCBI Assembly accession GCA_010125045.1). A core genome alignment was generated using Snippy v4.6.0 (default parameters; https://github.com/tseemann/snippy). Recombinant regions in the core genome alignment were masked using Gubbins v3.3.1 [47] with the options ‘--tree-builder iqtree-fast --first-tree-builder iqtree-fast’ (otherwise default parameters). snp-sites v2.5.1 [48] with ‘-c’ (otherwise default parameters) was used to extract variable sites in the recombination-masked alignment, resulting in a final recombination-free core SNP alignment. The final recombination-free core SNP alignment was used to calculate all-vs-all pairwise SNP distances using snp-dists v0.8.2 with default parameters (https://github.com/tseemann/snp-dists; **Dataset S7**). The above workflow was repeated to generate all-vs-all pairwise SNP distances for the 36 newly sequenced genomes alone, with publicly available genomes omitted (**Dataset S8**). Pairwise SNP distances were visualized using the R package pheatmap v1.0.12 (https://CRAN.R-project.org/package=pheatmap) with average linkage hierarchical clustering performed by hclust (from R package stats v4.3.2).

Phylogenetic inference was performed in IQ-TREE v2.2.5 [49], using the final recombination-free core SNP alignment for the full ST548 dataset as input and the following parameters: (i) the GTR+R nucleotide substitution model [50–52]; (ii) 1,000 ultrafast bootstrap replicates [53]; (iii) the number of constant sites in the alignment (obtained using snp-sites v2.5.1 with the ‘-C’ option), specified using the ‘-fconst’ parameter (i.e., ‘1078511,1182908,1166762,1071765’ for A, C, G, and T, respectively); (iv) ‘--seed’ was set to 1000. The resulting unrooted maximum likelihood (ML) phylogeny was imported into R v4.4.0, using the package ggtree v3.11.1 [54], and subsequently midpoint rooted using the ‘midpoint_root’ function in phytools v2.1-1 (referred to hereafter as the “midpoint-rooted ML phylogeny”; **Fig S3**) [55].

Two rooted, time-scaled phylogenies were additionally constructed using Rlsd2 v2.4.4 (https://github.com/tothuhien/Rlsd2) [56], with the unrooted ML phylogeny from IQ-TREE supplied as input, and tip dates corresponding to the year of isolation reported for each genome: (i) a fixed-rate ML phylogeny, with the options ‘seqLen=4708764,estimateRoot = “as”,confidenceInterval = 1000,givenRate = 1.27e-7’ (**Fig S4**) [14]; (ii) a LSD2- estimated-rate phylogeny, constructed as described in (i), but without supplying ‘givenRate = 1.27e-7’ to Rlsd2 (**Fig S5**). The LSD2-estimated-rate phylogeny indicated that a temporal signal may be present in the full ST548 dataset, although confidence intervals for the estimated rate and time to most recent common ancestor (tMRCA) were relatively large: LSD2 rate = 1.88268e-07 substitutions/site/year, confidence interval [6.9163e-08, 3.03092e-07]; tMRCA = 1886.88 [1685.09, 1936.07]. However, subsequent analysis of the unrooted ML phylogeny in TempEst v1.5.3 [57] did not reveal any evidence of a temporal signal (using the “best-fitting root” option: slope = 0, tMRCA = 2028, R-squared = 0.341) [57]. As such, Bayesian time-scaled phylogeny construction was not performed, and all three ML phylogenies (i.e., the midpoint-rooted, LSD2-fixed-rate, and LSD2-estimated-rate ML phylogenies) are provided (**10.5281/zenodo.15063662**).

### Core SNP calling and ML phylogeny construction for the high-AMR group

Additional ML phylogenies were constructed for a well-supported clade within the full ST548 dataset, which harbored many AMR determinants (i.e., the “high-AMR group”). Briefly, genome names for members of the high-AMR group were extracted from the larger ST548 phylogeny using the ‘mrcà function in the R package ape v5.7-1 (*n* = 108 high-AMR group genomes) [58]. The Snippy/Gubbins/snp-sites pipeline described above was used to identify core SNPs among all 108 high-AMR group genomes (see section “Core SNP calling and phylogeny construction”). The resulting recombination-free core SNPs were then used to build (i) midpoint-rooted (**Fig S6A**) and (ii) LSD2-estimated-rate ML phylogenies (**Fig S6B**) as described above (see section “Core SNP calling and phylogeny construction”). Evidence of a temporal signal was observed for the high-AMR group using both LSD2 and TempEst: LSD2 rate = 5.98136e-07 [3.97087e-07; 8.28845e- 07], tMRCA = 2009.89 [2004.52; 2012.52]; TempEst best-fitting root slope = 7.6933e-07, tMRCA = 2009.3, R-squared = 0.5046 for the unrooted ML phylogeny (**10.5281/zenodo.15063662**). As such, a Bayesian time-scaled phylogeny was constructed for the high-AMR group (described below).

### Bayesian phylogenetic reconstruction of the high-AMR group

BEAUti2 v2.7.6 and BEAST2 v2.7.6 [59] were used to construct a Bayesian time-scaled phylogeny of the high AMR group, using the high-AMR group recombination-free core SNP alignment as input and tip dates corresponding to each genome’s year of isolation (see section “Core SNP calling and ML phylogeny construction for the high-AMR group” above). A combination of (i) strict clock (starting clock rate = 1.27e-7 substitutions/site/year, with a lognormal prior on the clockRate parameter: mean = 2.77e- 7, standard deviation = 1.25, lower and upper bounds = [0, ∞]) [14] and (ii) coalescent Bayesian skyline model [60] were selected as the clock and population model, respectively, as these models have been identified previously as the optimal clock/population model combination for *S.* Minnesota [14]. The nucleotide substitution model was inferred using Bayesian model averaging (transitions and transversions split) with bModelTest v1.3.3 [61], and an ascertainment bias correction was used to correct for the lack of constant sites in the input core SNP alignment (number of constant sites obtained as described above in “Core SNP calling and ML phylogeny construction”; https://groups.google.com/g/beast-users/c/QfBHMOqImFE, accessed 13 March 2025).

Three independent BEAST2 runs were performed (using seeds 1000, 1001, and 1002), with chain lengths of 2 x 1e8 generations and parameters logged every 1e4 generations. The log file for each independent BEAST2 run was loaded into Tracer v1.7.2 to assess mixing (using 10% burn-in) and to confirm that effective sample size (ESS) values for all parameters were >200. The resulting log and tree files were combined using LogCombiner v2.7.6 with 10% burn-in. The final maximum clade credibility (MCC) tree was generated using TreeAnnotator v2.7.6 with common ancestor node heights. This MCC tree was loaded into R for visualisation as described above (see section “Core SNP calling and ML phylogeny construction”). The median effective population size over time was estimated using Bayesian skyline analysis in Tracer v1.7.2, using default parameters and the most recent sampling date set to ‘2022.5’.

To validate the observed evolutionary rate and tree height (i.e., to ensure that they were the result of a true temporal signal and not an artifact), a date randomization test was performed [62]. Briefly, sampling dates were randomized using the R package TipDatingBeast v1.1.0 [63], and the above BEAST2 workflow was performed for 10 independent sets of date-randomized samples. For these 10 randomized-date sample sets, models/parameters were identical to those described above, except for upper and lower clock-rate bounds on the “clockRate” parameter, which were set to 1e-3 and 1e-10, respectively (due to a ‘positive infinite posterior’ error, which occurred when bounds were not set, suggesting that our clock rate was escaping to extreme values in the date-randomized datasets). The date randomization test confirmed that there was no overlap between the 95% highest posterior density (HPD) intervals of the true clock rate/tree height and the clock rates/tree heights of the 10 independent date-randomized datasets (**Fig S7AB**). A prior-only run without any sequence data (‘sample from prior’ enabled in BEAUti2) was also performed to confirm minimal overlap in the resulting parameters between our prior-only analysis and our analysis with sequence data. Log files, XML files, and tree files for all BEAST2 runs described above are provided in the Zenodo repository (10.5281/zenodo.15063662).

### Identification of plasmid-mediated AMR genes

To determine which AMR genes were likely harbored on plasmids, the ‘end-to-end’ command in geNomad v1.7.1 [64] was used to classify all contigs in the full ST548 dataset as “chromosomal”, “plasmid”, or “viral” (see section “AMR determinant and plasmid replicon detection” above; **Dataset S9**). “Aggregated-classification” scores were compared between AMR gene-harboring (AMR+) contigs and contigs without AMR genes (AMR-; as determined by AMRFinderPlus) using the Kruskal-Wallis test in R (‘kruskal.test’ function). The “aggregated-classification” scores for the AMR+ contigs were then grouped by AMR gene and plotted in R using ggplot2 v3.5.1.

The plasmid database PLSDB version ‘2024_05_31_v2’ [65] was downloaded and converted into a BLAST v2.15.0 database using makeblastdb [66]. SeqKit v2.7.0 [67] was used to extract AMR+ contigs from the corresponding assemblies, and the contigs were then queried against the PLSDB BLAST database using megablast (‘blastn’ command with default parameters). Contigs that aligned to plasmid sequences in the PLSDB database at >95% coverage and >95% identity were considered to be candidate AMR gene-harboring plasmids (**Dataset S10**).

### Analysis of *bla*_CTX-M-8_-harboring contigs

Beta-lactamase *bla*_CTX-M-8_ was detected in several closely related ST548 genomes sequenced here (referred to hereafter as “ZA SNP cluster 0” genomes; described in detail in the “Results” section below). All *bla*_CTX-M-8_-harboring (*bla*_CTX-M-8_+) contigs underwent further investigation. Specifically, all *bla*_CTX-M-8_+ contigs matched a ∼90Kbp IncI pST113 plasmid from *Eschericha coli* (*E. coli*) in the PLSDB BLAST database with >95% coverage and >95% identity (NCBI Nucleotide accession CP134392.1, see section “Identification of plasmid-mediated AMR genes” above; **Dataset S10**). The accession number CP134392.1 was supplied as the query for a NCBI Nucleotide BLAST search against the ‘core_nt’ database, using ‘blastn’ as the selected program and the search limited to organism ‘Salmonella enterica (taxid:28901)’ (https://blast.ncbi.nlm.nih.gov/Blast.cgi, search conducted on 21 March 2025; **Dataset S11**) [68]. Aligned sequences for all hits were downloaded and underwent AMR determinant detection using AMRFinderPlus with the ‘-O Salmonella --plus’ options and database version 2024-05-02.2 (otherwise default parameters). This was done to identify similar plasmids in *Salmonella enterica*, and to determine if the hits also contained *bla*_CTX-M-8_. The only *bla*_CTX-M-8_+ hits were in two plasmids, both having >99% identity and >99% query coverage to the IncI pST113 plasmid from *E*. *coli*: (i) a ∼90Kbp plasmid previously identified in *S.* Mbandaka (NCBI Nucleotide accession CP146619.1) [69], and (ii) a recently uploaded ∼150Kbp plasmid sequence from *S.* Enteritidis (NCBI Nucleotide accession CP183502.1; **Dataset S12**). As the *S.* Mbandaka complete plasmid sequence was of similar size to the *E. coli* IncI plasmid (both ∼90Kbp) and associated with a publication [68], it was chosen as the reference sequence for further analysis.

### Reconstruction of *bla*_CTX-M-8_ + plasmid from ZA SNP cluster 0 genomes

Trimmed forward and reverse reads from all ZA SNP cluster 0 genomes (see section “Analysis of *bla*_CTX-M-8_-harboring contigs” above) were combined to create pooled forward and reverse reads. The pooled ZA SNP cluster 0 reads were then fed to the ‘plasmidspades’ command in SPAdes v4.0.0 [70], which was used to assemble plasmids among all pooled reads using the ‘--only-assembler’ parameter. The resulting ‘contigs.fastà file, the *S.* Mbandaka *bla*_CTX-M-8_+ IncI plasmid (NCBI Nucleotide accession CP146619.1), and the *E. coli bla*_CTX-M-8_+ IncI plasmid (NCBI Nucleotide accession CP134392.1) were annotated using Bakta v1.8.2 (see sections “AMR determinant and plasmid replicon detection” and “Analysis of *bla*_CTX-M-8_-harboring contigs” above). Using the corresponding Bakta .gbk files for these three plasmids, plasmid comparison and visualisation was performed using Clinker v0.0.31 [71] with the ‘-i = 0.5’ option. According to plasmidSPAdes, all ZA SNP cluster 0 contigs that mapped to CP146619.1 belonged to the same “plasmid component” (i.e., were predicted to be on the same plasmid). For the purposes of visualisation, contigs from the ZA SNP cluster 0 plasmidSPAdes assembly that did not align to the *S.* Mbandaka/*E. coli* reference IncI plasmids (i.e., assembled contigs assigned to a different “plasmid component”) were excluded from the figure. ABRicate v1.0.1 was used with the PlasmidFinder database to detect plasmid replicons in the plasmidSPAdes contigs as described above (see section “AMR determinant and plasmid replicon detection”; **Dataset S13**). We would like to note that while our PlasmidFinder results did not show the presence of IncI1 in any of the ZA SNP cluster 0 genomes individually (**Fig S3, Dataset S4,** see “AMR determinant and plasmid replicon detection” section above), IncI1 was detected in the pooled plasmidSPAdes assembly.

### Identification of *bla*_CTX-M–8_+ and other ZA ST548 genomes in the AllTheBacteria dataset

To assess the prevalence of *bla*_CTX-M-8_ in other bacterial lineages (e.g., other species, other *Salmonella* serotypes and sequence types, other ST548 genomes sequenced after our Enterobase data freeze), AMRFinderPlus results from AllTheBacteria (ATB) v0.2 (https://allthebacteria.readthedocs.io/en/latest/amr.html) were downloaded [72]. The ATB AMRFinderPlus results were then searched for “bla-CTX-M-8” (**Dataset S14**). All *bla*_CTX-M-8_-harboring ATB genomes classified as “*Salmonella enterica*” were downloaded (*n* = 64), and the resulting genomes underwent *in silico* seven-gene MLST using mlst v2.9 with default parameters. Sixteen genomes were assigned to ST548; these 16 genomes, plus the 14 ZA SNP cluster 0 genomes (see section “Analysis of *bla*_CTX-M-8_-harboring contigs” above), underwent all-vs-all pairwise SNP distance estimation as described above (see section “Core SNP calling and phylogeny construction”; **Dataset S15**). Metadata for all 354 *bla*_CTX-M-8_-harboring genomes from ATB were extracted using Entrez Direct v22.4 (**Dataset S16**) [73].

In addition to the 16 ST548 *bla*_CTX-M-8_-harboring genomes, we identified 10 other ST548 genomes from South Africa in the ATB dataset (all 10 were *bla*_CTX-M-8_-negative). These 10 genomes were present in Enterobase but did not have an associated NCBI assembly and were thus not included in our original data freeze. All-vs-all pairwise SNP distance estimation as described above (see section “Core SNP calling and phylogeny construction”) was performed with these 10 publicly available South African ST548 genomes and our 36 newly sequenced genomes. (**Dataset S17**).

## RESULTS

### Multiple *S.* Minnesota ST548 lineages can be detected in South African poultry imports of Brazilian origin

A total of 36 *Salmonella enterica* strains isolated from frozen raw poultry samples collected between 2020-2022 underwent WGS (see section “Sample isolation, DNA extraction, and WGS” above). While all 36 strains were isolated within South African borders, the frozen raw poultry from which each strain was isolated originated from Brazil. More specifically, (i) 11 out of 36 genomes (31%) were isolates originating from Brazilian poultry meat collected for regulatory compliance, at port-of-entry (POE); (ii) the remaining 25 isolates (69%) were from imported Brazilian poultry meat placed for sale in South African supermarkets (referred to hereafter as “**POE**” and “**supermarket**” genomes, respectively, with the total set of 36 genomes referred to as “**SEPI**” genomes [*<u>S</u>almonella <u>e</u>nterica* <u>P</u>oultry Imports]; **Table S1**).

Using *in silico* seven-gene MLST, all SEPI genomes belonged to ST548. *In silico* serotyping identified 35 genomes as serotype Minnesota (antigenic formula 21:b:e,n,x) by both SISTR and SeqSero2. One genome (strain 73, NCBI BioSample accession SAMN47166240) was assigned a serotype of “4:r:e,n,x” by SeqSero2 and “-:b:e,n,x” by SISTR. However, using core genome MLST (via SISTR), all SEPI genomes were identified as serotype Minnesota and will thus be referred to hereafter as *S.* Minnesota (see section “*In silico* serotyping and sequence typing” in the Methods above).

Despite belonging to the same ST, and despite sharing a similar origin (i.e., frozen raw poultry imported into South Africa from Brazil), considerable genomic diversity could be observed across SEPI genomes. ANI values calculated between SEPI genomes ranged from 99.84% - 100%. After removing recombination, the SEPI genomes differed by 0-85 core SNPs (mean 46, median 55; **Fig 1A**). The range of core SNP distances was not affected by the specific source of each genome, i.e., the core SNP distance range was approximately 0-85, even when comparing supermarket genomes only or POE genomes only (**Fig 1B**). Using average linkage hierarchical clustering of the pairwise core SNP distances, SEPI genomes could be partitioned into 7 SNP clusters at a threshold of 50 SNPs, to 21 SNP clusters at a threshold of 5 SNPs (**Fig 1C**). Altogether, these results indicate that multiple *S.* Minnesota ST548 lineages are present in our collection of isolates from Brazilian frozen poultry imports.

**Fig 1.**
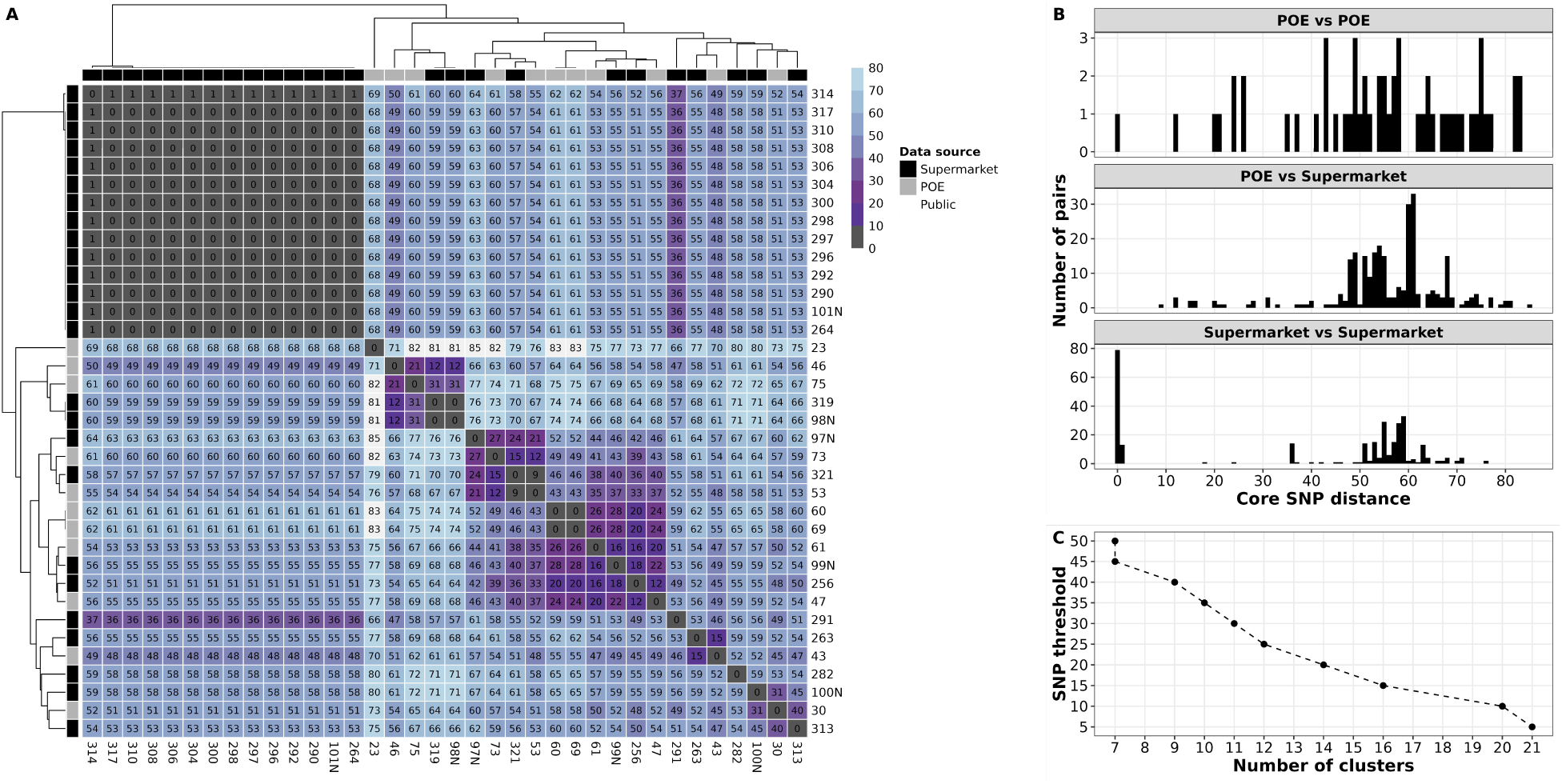
Core genome SNP distances show that multiple *S.* Minnesota ST548 lineages can be detected in Brazilian poultry imported into South Africa. (A) Heatmap showing pairwise core SNP distances calculated between the 36 ST548 genomes sequenced here (i.e., SEPI genomes). For the heatmap cells, darker shading indicates lower core SNP distances (i.e., more closely related genomes), while lighter shading indicates higher core SNP distances. Dendrograms were produced via average linkage hierarchical clustering of core SNP distances. Color strips above and to the left of the heatmap denote the source of isolation for each genome (“Data source”). POE - port-of-entry; Supermarket - imported poultry from supermarkets in South Africa. (B) Histogram of pairwise core SNP distances among the SEPI genomes separated by specific source of isolation. “POE vs POE” - comparisons within port-of-entry genomes only; “POE vs Supermarket” - comparisons between port-of-entry and supermarket genomes; “Supermarket vs Supermarket” - comparisons within supermarket genomes only. (C) Core SNP distance-based clustering of SEPI genomes. The X-axis shows the number of clusters at the corresponding core SNP threshold in the Y-axis. Clusters were obtained via average linkage hierarchical clustering (see “Core SNP calling and phylogeny construction” in the Methods above).

### *S.* Minnesota ST548 strains detected in South African supermarkets are closely related to strains isolated from Brazilian poultry imported into the United Kingdom

To gain insight into the evolution of South African ST548 within the context of the global ST548 population, we compared our 36 SEPI genomes to 228 high-quality, publicly available ST548 genomes (*n* = 264 total ST548 genomes; see section “Acquisition of publicly available WGS data and metadata” in the Methods above). Using Snippy, a total of 4,432 core SNP sites were identified among all 264 ST548 genomes, with SNP distances ranging from 0-322 (mean 160.5, median 193; **Fig S2A**).

For each SEPI genome sequenced here, we identified the corresponding publicly available genome with the lowest SNP distance. This approach identified 16 distinct publicly available genomes (ties were kept), which differed from SEPI genomes by 2-27 core SNPs (**Table S2**). Notably, 18 SEPI genomes (17 supermarket genomes) were 19- 27 SNPs apart from one Brazilian poultry-associated public genome (NCBI Assembly accession GCA_006209225.1). This suggests that nearly 70% of our supermarket genomes (17 out of 25 genomes) may share a recent common ancestor with a poultry-associated isolate from Brazil. In addition, 11 other ST548 strains from a Public Health England collection (NCBI BioProject accession PRJNA248792) were 5-50 core SNPs apart from all SEPI genomes. All 11 of these strains, though isolated in the UK, were associated with Brazilian imported poultry. This links SEPI isolates from South Africa with previously sequenced, poultry-associated strains from the UK, all of which share a common Brazilian origin. Collectively, these results suggest that while some South African ST548 genomes are closely related (<10 core SNPs) to previously sequenced food-associated genomes, others may represent previously unsampled lineages of similar origin (>20 core SNPs).

### *S.* Minnesota ST548 strains from South African poultry imports of Brazilian origin are confined to a largely AMR clade

A ML phylogeny constructed using the 4,432 core SNPs detected among all 264 ST548 genomes revealed a clear separation in line with geographical origin, AMR profile, and core SNP distances (**Fig 2, Fig S2AB**). One clade comprising 108 genomes of European, African, and Brazilian origin (ultrafast bootstrap support = 99%; referred to hereafter as the “high-AMR group”) was found to contain 88% of all AMR genes detected in our total ST548 sample set (937 of 1,063 AMR genes; **Fig 2, S3,S4, S5**). Further, all 108 genomes in the high-AMR group were predicted to be multidrug-resistant (MDR), i.e., harbored AMR genes conferring resistance to ≥3 antibiotic classes. The high-AMR group harbored 67% (70 of 104) of all European, 96% (25 of 26) of all African, and 59% (13 of 22) of all South American ST548 genomes (**Fig S5, Fig S6**). Notably, the high-AMR group also included all 36 of our SEPI genomes. The remaining genomes from Europe, Africa, and South America (*n* = 40), along with all North American and Asian genomes (*n* = 106 and 7, respectively) belonged to other clades within the ST548 phylogeny and contained only ∼12% of all AMR genes detected (126 of 1,063 AMR genes; *n* = 154 total genomes, referred to hereafter as the “low-AMR group”). A predicted MDR phenotype was observed among low-AMR group genomes only sporadically (13 of 154 low-AMR group genomes, 8.4%; **Fig S5**).

**Fig 2.**
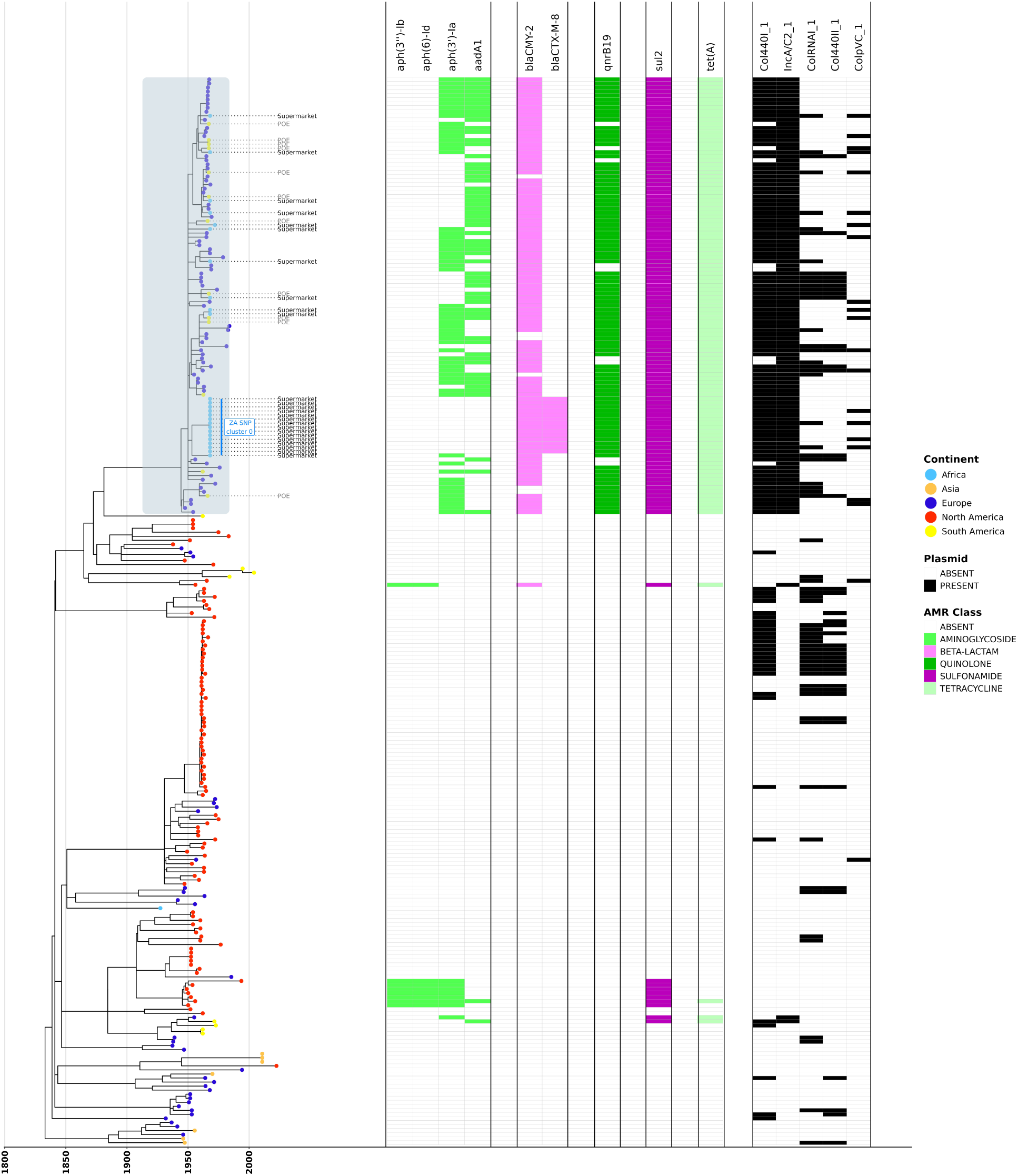
LSD2-estimated-rate maximum likelihood (ML) phylogeny of *S.* Minnesota ST548 shows that the majority of AMR genes are confined to a single clade (*n* = 264; 36 SEPI genomes, plus 228 high-quality, publicly available *S.* Minnesota ST548 genomes). The “high-AMR group” is denoted in the phylogeny by light blue shading. Tree tips for the 36 SEPI genomes are annotated with the isolation source (“POE” for strains isolated at port-of-entry; “Supermarket” for poultry samples from supermarkets in South Africa). Unlabelled tips denote publicly available genomes from Enterobase. Tips are colored by continent of isolation (“POE” and “Supermarket” genomes sequenced in this study were classified as “South American” and “African” isolates, respectively). Heatmaps to the right of the phylogeny denote (from left to right): (i) presence/absence of AMR genes detected by AMRFinderPlus, colored by AMR class; (ii) presence/absence of plasmid replicons (detected using ABRicate/PlasmidFinder). For readability, only select AMR genes and plasmid replicons are shown (see Fig S5 for the full version of this figure). The ML phylogeny was constructed using IQ-TREE, and the evolutionary rate was estimated by LSD2, with branch lengths reported in years (to view phylogenies constructed using different rooting/scaling methods, see Fig S3, S4).

The separation between the high- and low-AMR groups was also evident when ST548 genomes were clustered by accessory gene presence/absence (i.e., genes present in >5% and <95% of ST548 per Panaroo). As was the case with the core genome ML phylogeny, all SEPI genomes clustered with publicly available genomes from the high- AMR group (**Fig 3**). Interestingly, two genomes from the low-AMR group clustered with high-AMR group genomes (NCBI Assembly accessions GCA_011578495.1 and GCA_022318025.1; **Fig 3**). These two genomes belonged to the small group of sporadic AMR+ isolates in the low-AMR group and were positive for aminoglycoside, sulfonamide, and tetracycline resistance genes (**Fig S5**).

**Fig 3.**
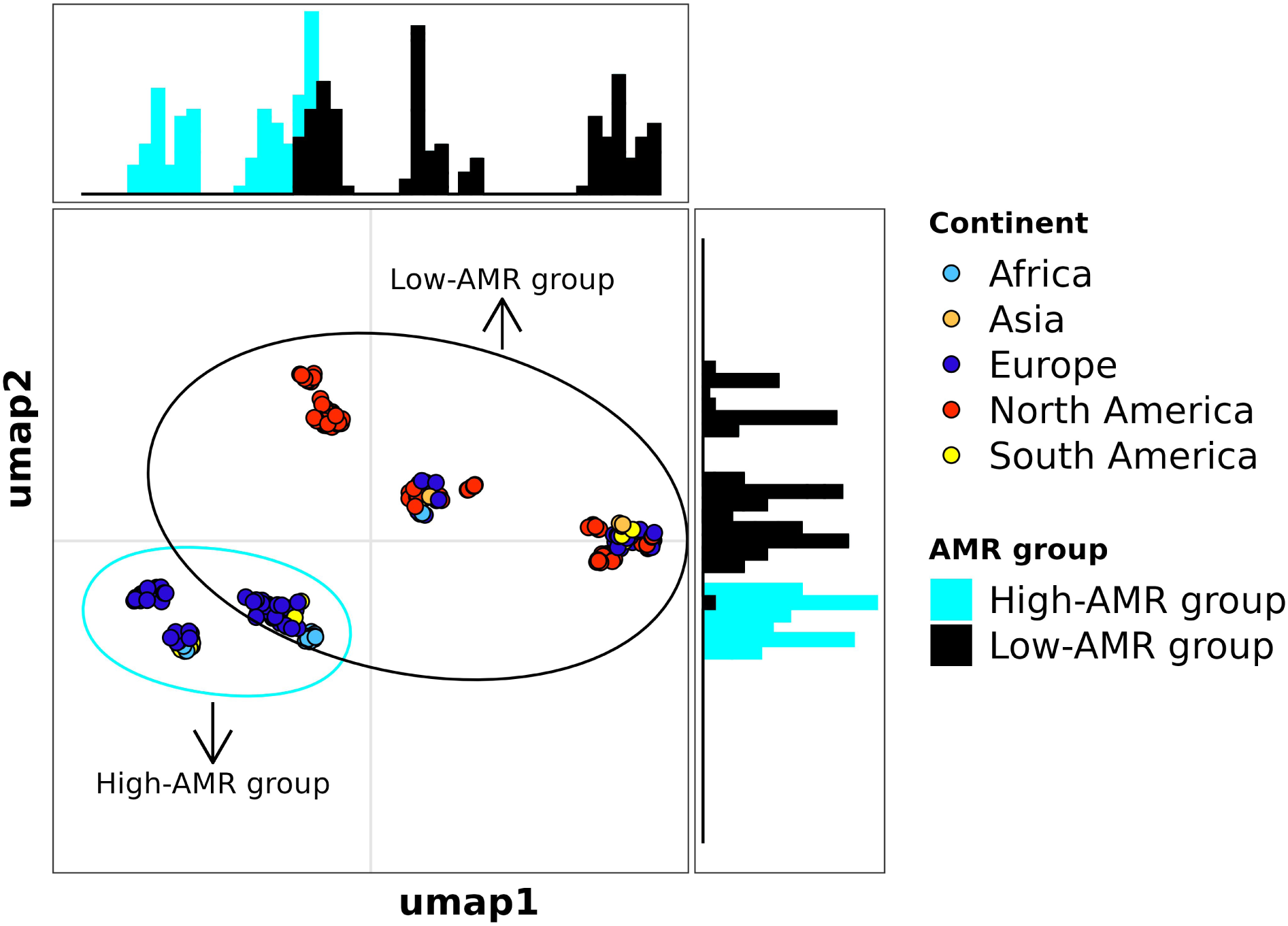
UMAP constructed using accessory gene presence/absence reveals separation between high-AMR and low-AMR groups. The UMAP was constructed from a presence/absence matrix of intermediate genes detected among 264 *S.* Minnesota ST548 genomes (i.e., genes present in >5% and < 95% of the population according to Panaroo). Each point represents a genome, colored by the reported continent of isolation. Ellipses encompass all genomes from the high-AMR group (cyan) or low-AMR group (black). Histograms along the X and Y axes of the plot represent the distribution of high-AMR group and low-AMR group genomes along the corresponding axis in the UMAP.

To further investigate the emergence of the high-AMR group, we constructed a Bayesian time-scaled phylogeny of the 108 high-AMR group genomes (see section “Bayesian phylogenetic reconstruction of the high-AMR group” in the Methods above). The evolutionary rate for the high-AMR group was estimated to be 6.262e-07 substitutions/site/year (95% HPD [5.428e-07, 7.102e-07]), with a tMRCA of 2010.485 (95% HPD [2009.02, 2011.83]; **Fig 4**). Bayesian skyline analysis estimated that the effective population size of the high-AMR group underwent an increase in ∼2013 and then subsequently a decrease post 2020 (**Fig S7C**). However, such sharp population changes can be an artifact of sampling and population structure as demonstrated previously [74, 75].

**Fig 4.**
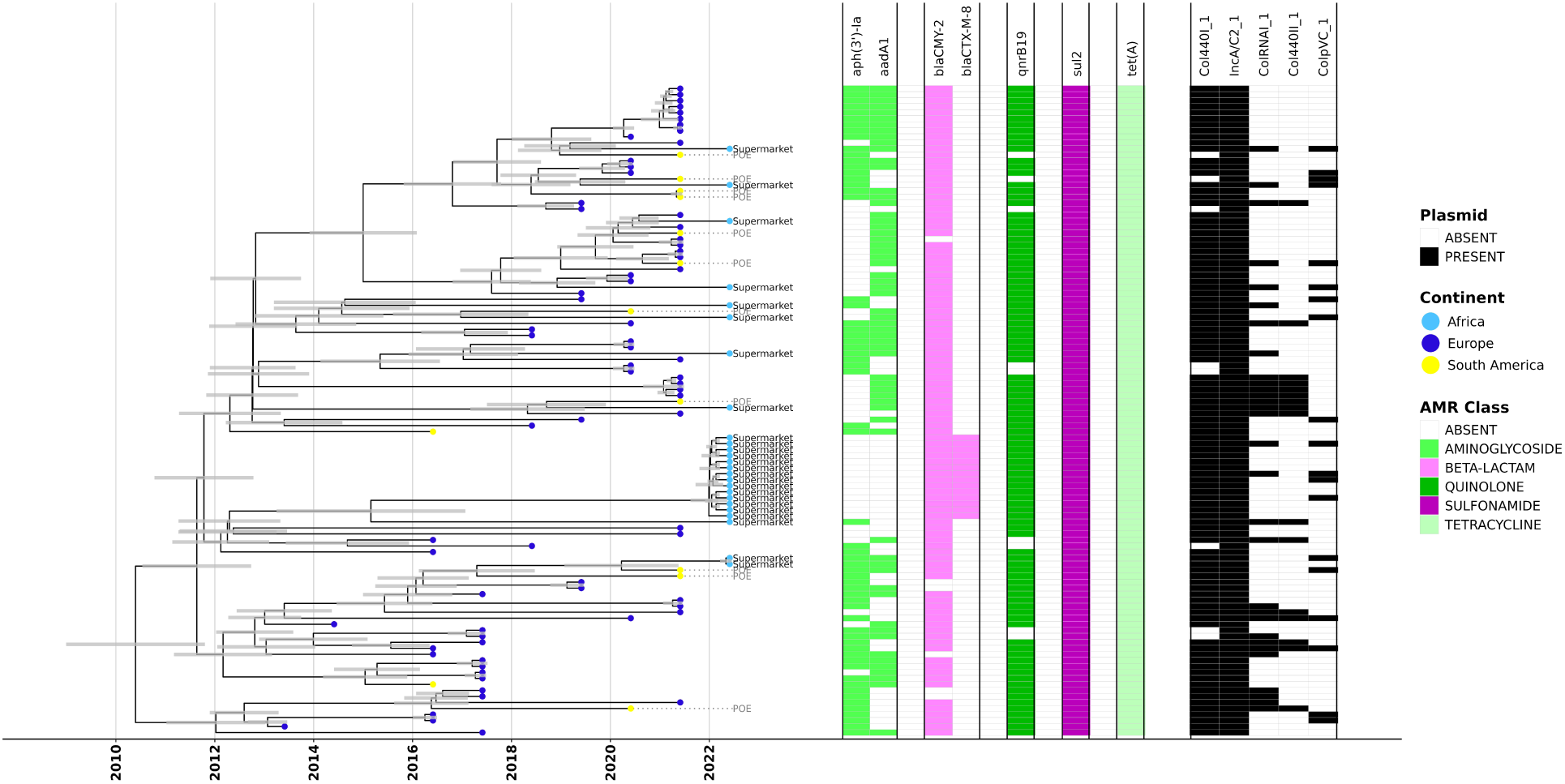
Bayesian phylogenetic reconstruction shows recent emergence of the high- AMR group. (comprising 108 isolates, including all 36 SEPI genomes). The X-axis denotes time in years, and the grey node bars represent 95% highest posterior density (HPD) intervals for node heights. Tree tips for the 36 SEPI genomes are annotated with the isolation source (“POE” for strains isolated at port-of-entry; “Supermarket” for poultry samples from supermarkets in South Africa). Unlabelled tips denote publicly available genomes from Enterobase. Tree tips are colored by the continent of isolation (“POE” and “Supermarket” isolates sequenced in this study were classified as “South American” and “African” isolates, respectively). Heatmaps to the right of the phylogeny denote (from left to right): (i) presence/absence of AMR genes detected by AMRFinderPlus, colored by AMR class; (ii) presence/absence of plasmid replicons (detected using ABRicate/PlasmidFinder). Only select AMR genes and plasmid replicons are shown (to view the full phylogeny, see Fig S6).

### Antimicrobial resistance genes in *S.* Minnesota ST548 are largely plasmid- associated

To investigate whether AMR genes from the high-AMR group were plasmid-associated, geNomad was used to predict the origin (chromosomal, plasmid, or viral) of assembled contigs in the full ST548 set (**Fig 5**). geNomad predicted plasmid scores for contigs harboring AMR genes (AMR+ contigs) were then compared to contigs without AMR genes (AMR- contigs) as detected by AMRFinderPlus. Overall, plasmid scores were significantly greater for AMR+ contigs compared to AMR- contigs (*P* < 2.22e-16, Mann Whitney *U* test, **Fig S8**). Using blastn, each AMR+ contig aligned to ≥1 plasmid in PLSDB with >95% coverage and >95% identity (see “Identification of plasmid-mediated AMR genes” in the Methods above). Contigs harboring *bla*_CMY-2_, *tet*(A), and *sul2* aligned to >400 distinct ∼100 Kbp plasmids, each with coverage and identity >95%. Moreover, 70 of these ∼100Kbp plasmids were found to harbor all three AMR genes. This suggests that MDR-conferring megaplasmids are in circulation in this lineage, as has been noted previously [17]. While we were unable to pinpoint the exact plasmid due to the fragmented nature of short-read assemblies, we identified reference sequences for potential candidates (NCBI Nucleotide accessions NZ_CP012923.1, NZ_CP060509.1, or NZ_CP080427.1).

**Fig 5.**
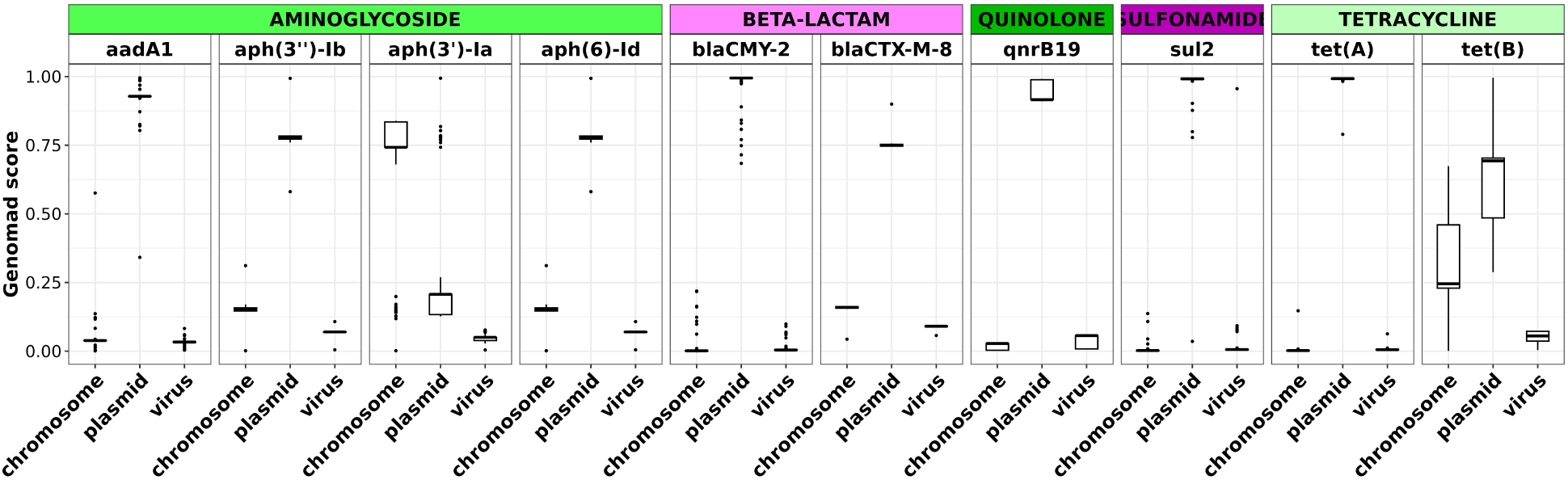
geNomad scores for ST548 AMR+ contigs show that most AMR genes are plasmid-associated. Boxplots showing geNomad aggregated classification scores for AMR gene-harboring (AMR+) contigs present among 264 *S.* Minnesota ST548 genomes. Black horizontal lines within each boxplot denote the median geNomad score, and boxes show the interquartile range. Whiskers represent values up to 1.5x the first (lower) or third (higher) quartile, with black points showing outliers beyond the whiskers’ range. Each facet shows the geNomad chromosome, plasmid, and phage score for the corresponding AMR gene. AMR genes are grouped by AMR class.

### A *bla*_CTX-M-8_-harboring plasmid from *E. coli* has entered South Africa via Brazilian poultry imports

Among the 36 SEPI genomes sequenced here, the largest SNP cluster consisted of 14 isolates, all 0 core SNPs apart (referred to hereafter as “ZA SNP cluster 0”; **Fig 2 and Fig S1**). All 14 ZA SNP cluster 0 genomes were isolated from poultry meat sold in supermarkets in South Africa (**Fig 2**). A unique feature of genomes within ZA SNP cluster 0 was that they harbored *bla*_CTX-M-8_, which was otherwise absent in our full *S.* Minnesota ST548 dataset (**Fig 2**). This was in addition to *bla*_CMY-2_, *sul2*, and *tet*(A), which were, as stated previously, potentially carried by a megaplasmid (see section “Antimicrobial resistance genes in *S.* Minnesota ST548 are largely plasmid associated” above).

To assess the prevalence of *bla*_CTX-M-8_ in other *Salmonella enterica* lineages, we searched a catalog of AMR determinants detected in >1.9 million bacterial genomes (i.e., AMRFinderPlus results from the AllTheBacteria [ATB] dataset) [72]. Out of 644,699 ATB *Salmonella enterica* genomes, only 64 possessed *bla*_CTX-M-8_ (0.01%), suggesting that *bla*_CTX-M-8_ is not common in *Salmonella enterica* (see “Identification of *bla*_CTX-M–8_+ and other ZA ST548 genomes in the AllTheBacteria dataset” in the Methods above). Out of these 64 *bla*_CTX-M-8_-harboring (*bla*_CTX-M-8_+) ATB *Salmonella enterica* genomes, 16 belonged to ST548 (25%). These 16 *bla*_CTX-M-8_+ ST548 ATB genomes were not included in our initial dataset because there was no associated GenBank assembly available at the time of our data freeze, or the existing assembly did not pass our quality filters (see section “Acquisition of publicly available WGS data and metadata” in the Methods above). Further investigation of these 16 ATB genomes revealed all 16 (100%) differed from ZA SNP cluster 0 by >50 core SNPs (**Fig S8**), indicating that ZA SNP cluster 0 may not share a common source with any of the *bla*_CTX-M-8_+ ST548 genomes from ATB. Further, two of these 16 *bla*_CTX-M-8_+ ST548 ATB genomes (12.5%) differed from ZA SNP cluster 0 by >250 core SNPs, meaning they are likely not part of the high-AMR group itself (**Fig S9, S2B**). Interestingly, one of these 16 *bla*_CTX-M-8_+ ST548 ATB genomes was reported to be from South Africa (NCBI BioSample accession SAMEA14452848), isolated in 2022 from animal meat along with 10 other *bla*_CTX-M-8_- samples. Each of these 11 South African samples were 23-65 core SNPs apart from at least one SEPI genome sequenced here (**Table S3**). From the remaining 15 *bla*_CTX-M-8_+ ST548 ATB genomes, five were from Brazilian poultry, isolated between 2016 and 2022; nine were reported to be isolated from “food” in Portugal and the UK. The one remaining sample from the UK was reported to be isolated from a human (NCBI BioSample accession SAMN30444884). Taken together, our results suggest that, while *bla*_CTX-M-8_ is relatively rare in *Salmonella enterica*, ST548 has been subjected to multiple independent introductions of *bla*_CTX-M-8_.

All ST548 *bla*_CTX-M-8_+ contigs (i.e., from ZA SNP cluster 0, as well as the 16 ST548 genomes from ATB) matched an IncI pST113 plasmid from *E. coli* with >95% coverage and >95% identity (NCBI Nucleotide accession CP134392.1). A similar *bla*_CTX-M-8_+ plasmid has previously been reported in *Salmonella enterica* serotype Mbandaka (*S.* Mbandaka; NCBI Nucleotide accession CP146619.1) [69]. In an attempt to assemble the complete plasmid, we pooled all reads from the ZA SNP cluster 0 genomes and ran plasmidSPAdes. We then annotated the resulting contigs with Bakta and compared the resulting gene clusters with the *S.* Mbandaka and *E. coli bla*_CTX-M-8_+ IncI plasmids using Clinker (**Fig 6**). This showed that, in addition to an existing MDR-conferring megaplasmid commonly found in the high-AMR group of *S.* Minnesota ST548, 14 SEPI genomes have an additional plasmid with *bla*_CTX-M-8_. Using AMRFinderPlus results from ATB, we found a total of 354 genomes with *bla*_CTX-M-8_; of these, 248 were detected in *E. coli* (70.1%), including the earliest reported *bla*_CTX-M-8_+ sample in the ATB dataset (NCBI BioSample accession SAMN33422834, isolated in 2005 from a rectal swab of a dog in the United Arab Emirates; see “Identification of *bla*_CTX-M–8_+ and other ST548 genomes in the AllTheBacteria dataset” in the Methods above). Overall, this suggests that the *bla*_CTX-M-8_+ IncI plasmid was potentially transferred to ST548 from *E. coli*.

**Fig 6.**
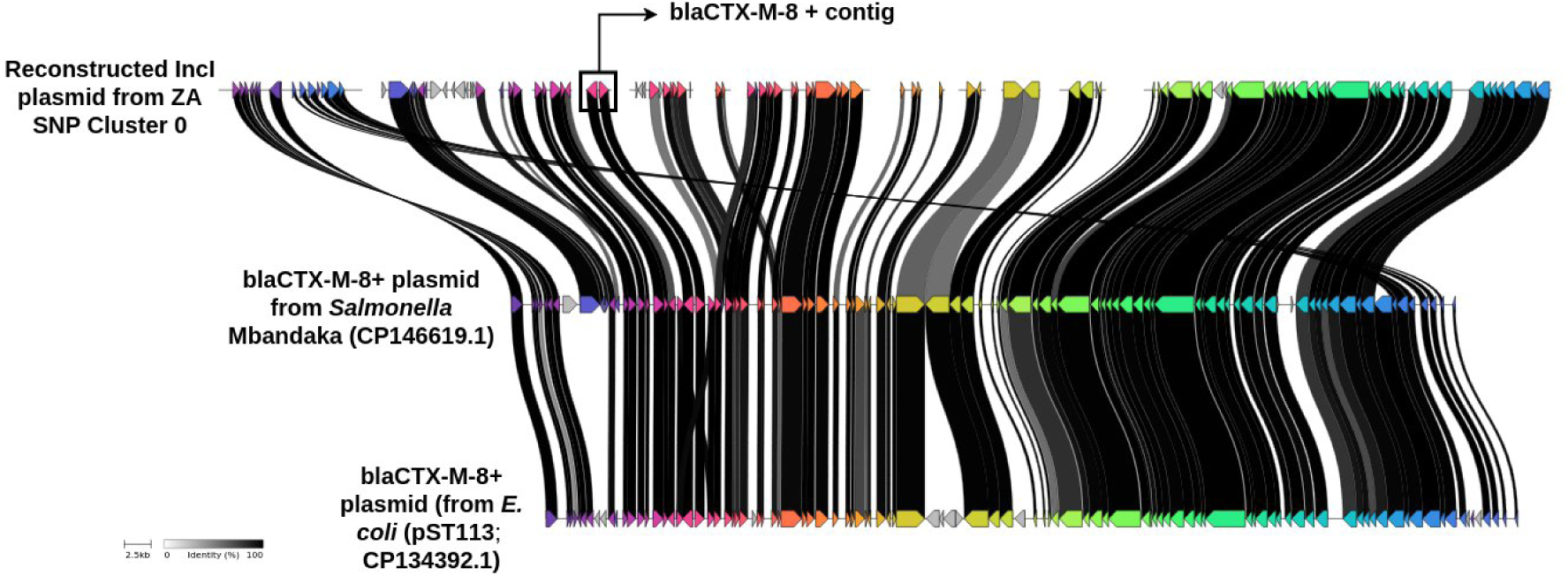
*bla*_CTX-M-8_-harboring contigs from ZA SNP cluster 0 are highly similar to an IncI1 plasmid from *S.* Mbandaka and *E. coli*. Plot showing the synteny and relatedness of *bla*_CTX-M-8_-harboring IncI1 plasmids detected in genomes sequenced here (i.e., ZA SNP cluster 0) and the most closely related plasmids, detected in *S.* Mbandaka (CP146619.1) and *E. coli* (CP134392.1). The bands connecting each gene (arrows) indicate amino acid sequence identity, with darker bands indicating higher identities. Sequences with <50% identity are not connected. The color of each gene represents the functional annotation group.

## DISCUSSION

### Port-of-entry genomic surveillance provides insight into the global dissemination of pathogens via agro-food trade

Bacterial pathogens endemic to a given geographical region can be exposed to unique, local selection pressures, which shape their evolution and population structure (e.g., exposure to antimicrobials in agricultural, environmental, or clinical settings, dictated by local regulations or the lack thereof) [76–79]. For foodborne zoonotic pathogens like *Salmonella enterica*, the global agro-food trade serves as a mechanism by which endemic pathogens can be disseminated to other world regions, potentially infecting humans and/or animals [4, 5, 80–84]. As such, many countries routinely test agro-food imports for the presence of pathogens, with the goal of preventing contaminated imports from entering domestic food systems [84–86]. In South Africa, under the Meat Safety Act, 2000 (Act No.40 of 2000) and related Veterinary Procedural Notifice (VPN), microbiological testing is required for compliance monitoring of both domestic and imported meat products. Upon arrival and during storage, random samples are collected from selected consignment packages and sent to approved laboratories for bacteriological testing, including *Salmonella* detection [24]. Since 2018, VPN 56 mandates that all *Salmonella* strains isolated from meat samples in South Africa be forwarded to the ARC: OVR- General Bacteriology Laboratory for storage and molecular typing monitoring [87].

Here, we used WGS to characterize 36 *S.* Minnesota ST548 strains isolated in South Africa, from imported poultry of Brazilian origin. In addition to querying ST548 strains from poultry meat sold in supermarkets, our sampling efforts included ST548 strains isolated from poultry meat consignments held at port-of-entry. Notably, our WGS data showed that *S*. Minnesota ST548 strains detected in South African supermarkets were closely related to strains isolated in poultry meat exported from Brazil to the UK. All 36 of our isolates were confined to a well-supported clade within the ST548 phylogeny (i.e., the “high-AMR group”), which contained at least eleven other isolates known to be associated with Brazilian poultry imports. These eleven isolates were reported in a previous study from the UK, in which *S.* Minnesota genomes from UK poultry meat imports of Brazilian origin formed a monophyletic clade with genomes from Brazil [4]. Further, seven of our genomes were highly similar to genomes from the UK (≤10 core SNPs; **Table S2**), indicating that some (but not all) European and African imports may share a common source. We also found a set of 11 South African food-associated ST548 genomes from ATB, which were 23-65 core SNPs apart from at least one of our SEPI genomes (**Table S3**).

Overall, currently available WGS data suggests that several *S.* Minnesota ST548 lineages have been introduced into South Africa through Brazilian poultry imports. The results showcase the value of rich geographic metadata in pathogen surveillance studies, particularly metadata conveying the provenance of imported foods. As shown here and elsewhere [21, 22, 24, 88, 89], foodborne pathogens isolated at POE, from a known exporter, are particularly useful, as they can capture pathogen migration events between geographic regions with known directionality.

### Existing publicly available WGS data cannot link *S.* Minnesota ST548 from Brazilian poultry imports to South African human clinical cases

Some importers have expressed concern regarding *Salmonella* in Brazilian meat and poultry [4, 25, 90, 91]. However, the increasing prevalence of *S.* Minnesota in poultry may not necessarily translate to an increase in human salmonellosis cases. For example, in the UK, there was no observable increase in human clinical cases caused by *S.* Minnesota, despite the increasing prevalence of *S.* Minnesota in Brazilian poultry, and the few observed human clinical cases could largely be explained by recent international travel [4]. The European Centre for Disease Prevention and Control (ECDC) infectious disease surveillance atlas shows only 354 total reported cases of human salmonellosis caused by *S*. Minnesota from 2007-2023 in the EU (UK not included after 2019). Another common Brazilian poultry-associated serotype, *S.* Heidelberg, showed 1,790 cases in the same time period, though with a decreasing trend. These figures are dwarfed by serotype *S.* Enteritidis, which showed 33,088 cases in the EU in 2023 alone (https://atlas.ecdc.europa.eu/public/index.aspx). Overall, these data suggest that *S.* Minnesota may not be a significant threat to human health compared to other *Salmonella* serotypes in Europe.

Here, we found that all *S.* Minnesota strains isolated in South Africa are relatively closely related to food/poultry associated strains from Brazil or the UK (<65 core SNPs; **Fig S2A, Table S2, Table S3**). Therefore, it is possible that Brazilian *S.* Minnesota ST548 also does not impose a significant disease burden on South African consumers, as evidence indicates for the UK and EU [4]. However, a lack of links to clinical cases could rather be the result of data gaps. Despite imposing a disproportionately high burden of illness on sub-Saharan Africa [92, 93], *Salmonella* WGS efforts are largely concentrated in world regions where the burden of salmonellosis is lower [92, 94, 95]. For example, of the 228 Enterobase *Salmonella* ST548 genomes included in our dataset, 188 (∼82%) originated from strains isolated in the US or the UK. While African-led pathogen surveillance efforts are producing an unprecedented amount of data, African WGS data generators can still face major hurdles when sharing, publishing, and/or disseminating pathogen WGS (meta)data [96]. Thus, the lack of observed links between imported *S.* Minnesota and human clinical cases in South Africa could be due to the lack of publicly available WGS data from *S.* Minnesota strains in South Africa (e.g., from human clinical cases, as well as domestic animals, foods, and environmental sources). Overall, the UK experience with *S.* Minnesota, both in terms of surveillance and disease burden, cannot be applied directly to South Africa, and our study highlights the importance of local pathogen surveillance initiatives, beyond those conducted in Europe and North America.

### Multiple *bla*_CTX-M-8_-harboring *S.* Minnesota ST548 lineages have been disseminated via international poultry trade

Antimicrobial use in livestock and food production environments can select for AMR foodborne zoonotic pathogens, which can be subsequently disseminated via international trade [97–100]. Consequently, farm management and antimicrobial use practices in a single country, region, or even farm can impact human and animal health across the globe [100, 101]. Here, we identified a closely related *S.* Minnesota ST548 lineage (i.e., ZA SNP cluster 0, differing by 0 core SNPs), which had acquired *bla*_CTX-M-8_. *bla*_CTX-M-8_ was published as a novel CTX-M beta-lactamase in 2000 [102], when it was identified in three strains of three species (*Enterobacter cloacae*, *Enterobacter aerogenes*, and *Citrobacter amalonaticus*) isolated in 1996-1997 from intensive care unit patients in Rio de Janeiro, Brazil. Since then, studies in Brazil and other South American countries have noted that *bla*_CTX-M-8_ appears to be relatively common among extended-spectrum beta-lactamase (ESBL)-producing *E. coli* in the region [103, 104]. However, *bla*_CTX-M-8_ has been detected in numerous other countries, in a range of hosts (e.g., *E. coli*, *Shigella* spp., *Enterobacter* spp., *Klebsiella* spp., *Salmonella enterica*), isolated not only from human clinical cases, but apparently healthy humans (e.g., working as food handlers), animals/animal products (e.g., chicken meat, chickens, cattle), wastewater, and wildlife (e.g., gulls, wild birds) as well [103–115]. Most notably within the context of this study, *bla*_CTX-M-8_ has previously been detected in *E. coli* isolated from Brazilian poultry products imported into Japan [105, 116], indicating that this AMR gene has previously been disseminated intercontinentally via Brazilian poultry.

In addition to ZA SNP cluster 0, we identified an additional 16 *bla*_CTX-M-8_-harboring *S.* Minnesota ST548 genomes, isolated between 2017-2022 (three samples had unreported isolation dates) from 4 different countries (i.e., Brazil, Portugal, South Africa and the UK; two samples had unreported isolation locations). Notably, two of these *bla*_CTX-M-8_- harboring *S.* Minnesota ST548 genomes (NCBI BioSample accessions SAMN30444884, SAMEA12320398), one from Brazil and one from the UK, were 91 core SNPs apart and >290 core SNPs apart from ZA SNP cluster 0. (**Fig S9**). This indicates that multiple independent acquisition events are responsible for the emergence of *bla*_CTX-M-8_ in ST548. Previous studies have identified *bla*_CTX-M-8_ on plasmids, including IncI1 plasmids harbored by *E. coli* [103, 105, 113]. Notably, *bla*_CTX-M-8_-harboring IncI1 plasmids have been detected across multiple *E. coli* STs [103, 105, 113], indicating that multiple *E. coli* lineages have acquired this AMR gene via IncI1 plasmids. Further, a *bla*_CTX-M-8_-harboring IncI1 plasmid was recently described in a *Salmonella* Mbandaka strain (isolated in 2022 from a broiler farm environment in Poland) [69].

Overall, *bla*_CTX-M-8_ can be acquired within and across species boundaries via IncI1 plasmids. Based on the genomic analyses conducted here, we hypothesize that ZA SNP cluster 0 acquired *bla*_CTX-M-8_ recently, within Brazil, via an *E. coli* IncI1 plasmid, and was subsequently disseminated to South Africa via imported poultry. However, whether *bla*_CTX-M-8_-harboring *S.* Minnesota imposes a significant burden on humans and animals in South Africa remains a mystery. Future WGS efforts targeting *S.* Minnesota from human and non-human sources (e.g., poultry flocks, farm environments, poultry products) in South Africa are needed. We anticipate that long-read sequencing methods in particular will provide insight into AMR dynamics within *S.* Minnesota. This, combined with continued surveillance of imported foods at port-of-entry, will allow for improved risk evaluation of imported foods in South Africa.

## Data Availability

All data produced are available online at https://github.com/VishnuRaghuram94/SEPI and https://zenodo.org/records/15063662

https://github.com/VishnuRaghuram94/SEPI

## Acknowledgments

Computation was enabled by resources provided by (i) High Performance Computing Center North (HPC2N; Umeå University, Umeå, Sweden), and (ii) the National Academic Infrastructure for Supercomputing in Sweden (NAISS), partially funded by the Swedish Research Council through grant agreement no. 2022-06725. The bacterial isolates sequenced in this study were supplied by the Agricultural Research Council – Onderstepoort Veterinary Research.

## Author contributions

V Raghuram performed all computational analyses with input from TM, V Ramnath, HG, JB, and LMC. IM and LMC conceptualized and funded the study. V Raghuram, LMC, KM, and IM co-wrote the manuscript with input from all co-authors.

