## Supplemental Figures S1 - S9 and Supplemental table S1 - S3 legends for "Genomic surveillance of *Salmonella enterica* serotype Minnesota strains from poultry products imported into South Africa"

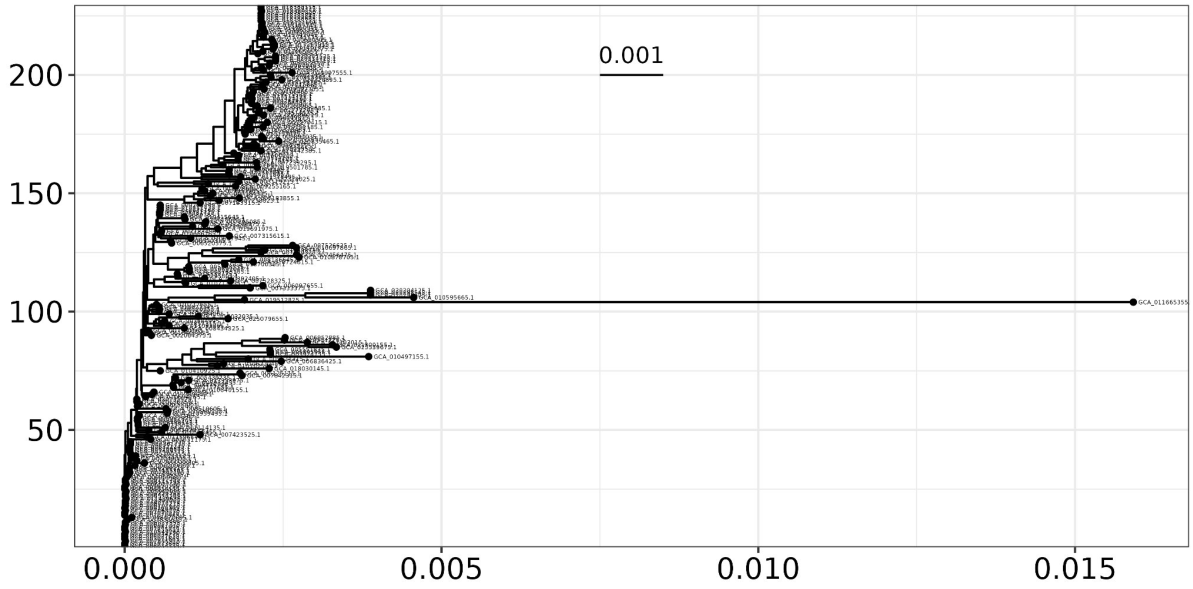


**Fig S1**: Mash distance-based tree of 229 curated publicly available *S*. Minnesota ST548 genomes revealed a single outlier sample (GCA_011665355.1) which was then removed from the dataset before further analyses.

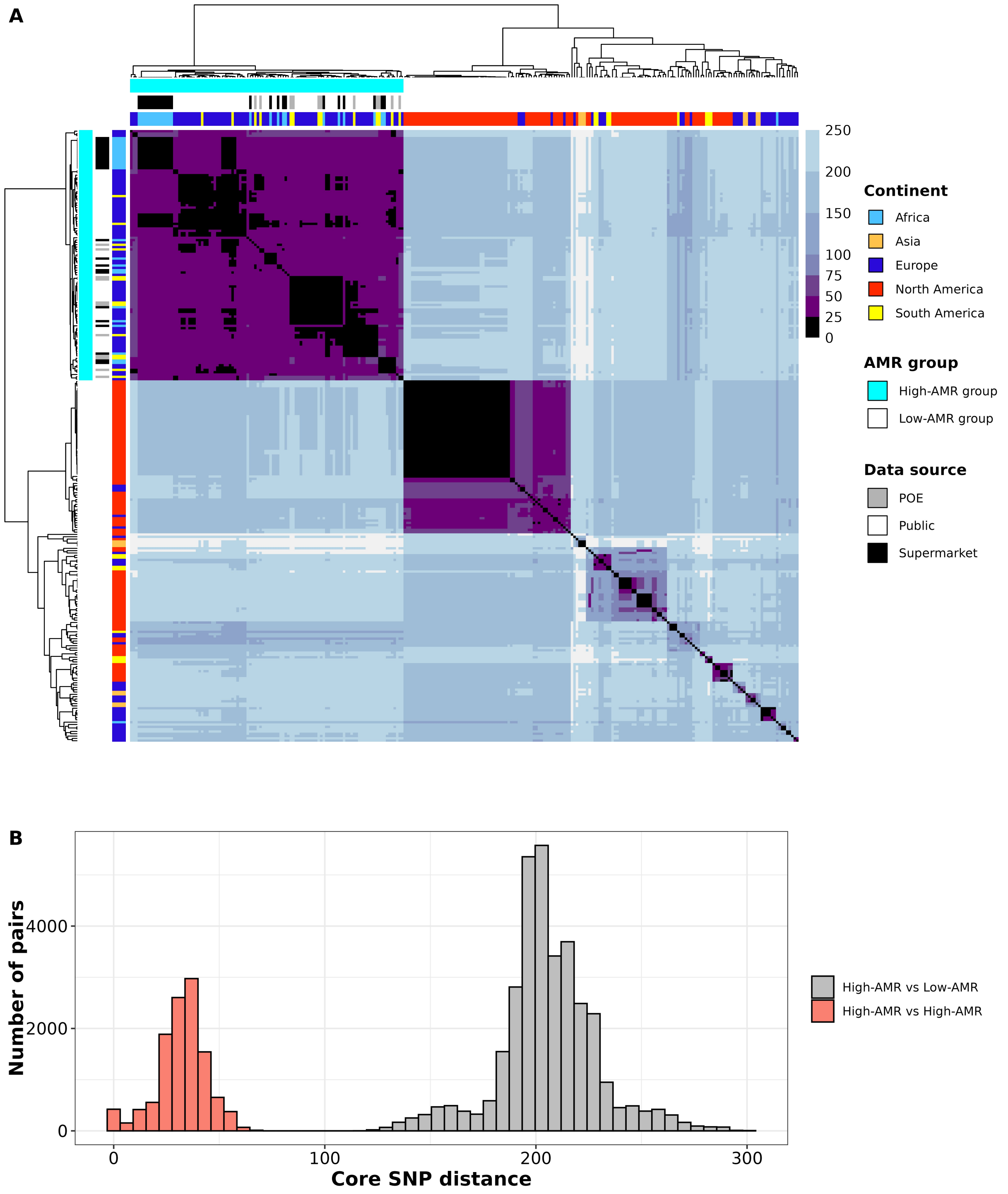


**Fig S2: (A)** Heatmap showing pairwise core genome SNP distance (lighter = higher SNP distance) for all 264 ST548 isolates. Heatmap is annotated by 1) continent of isolation; 2) source of the data (POE - Port of entry, Supermarket - poultry samples from supermarkets in South Africa, Public data - ST548 genomes uploaded to Enterobase) and 3) AMR prevalence (high-AMR group/low-AMR group). **(B)** Histogram of pairwise core SNP distances between only genomes within the high-AMR group (red bars) or between the high-AMR group and low-AMR group (grey bars).

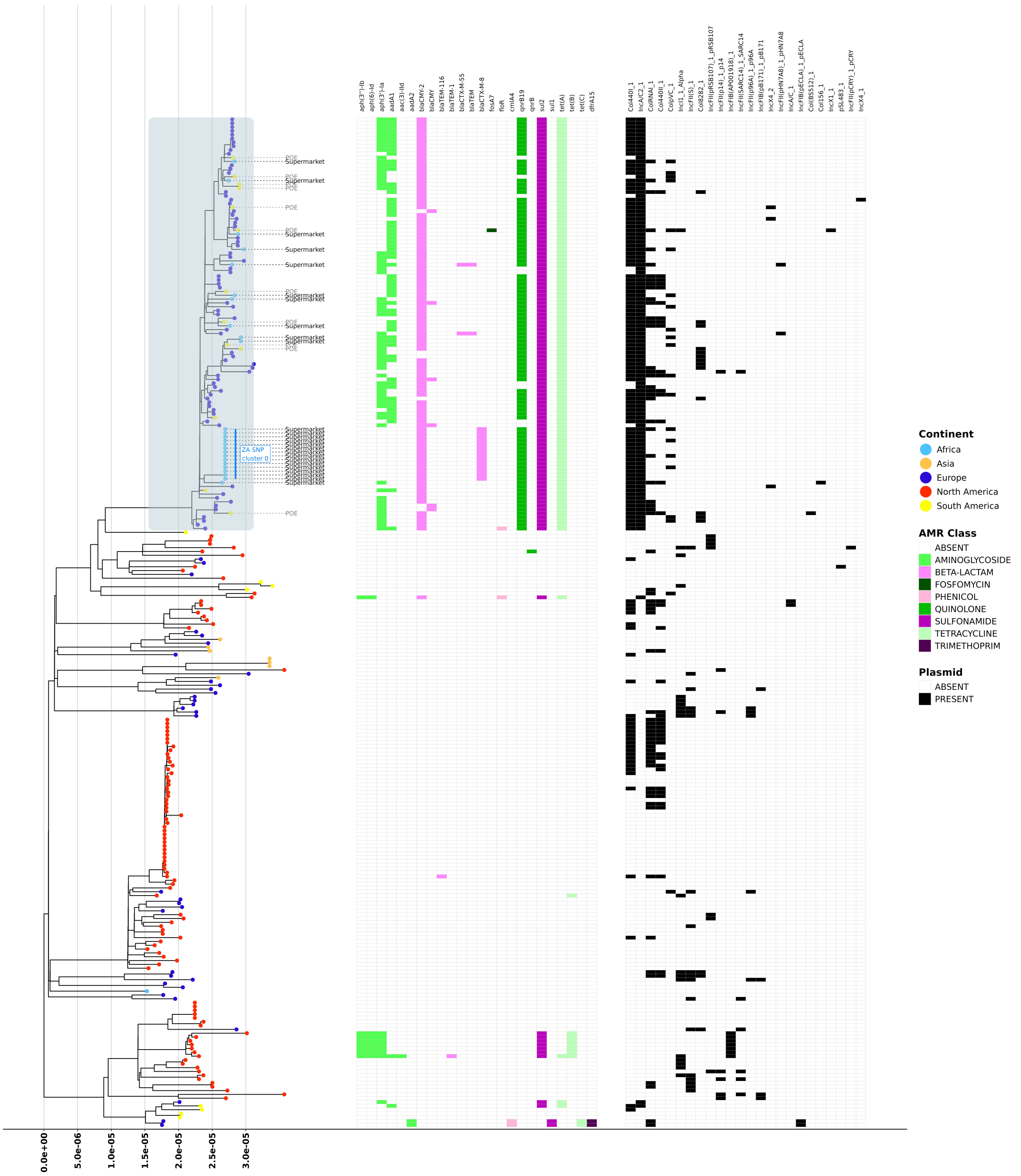


**Fig S3:** Midpoint rooted maximum-likelihood (ML) phylogenetic tree constructed using IQ-TREE comprising all 264 ST548 isolates.Tree tips are coloured by continent of isolation. Heatmap corresponding to tree tips indicate the source of the data (POE - Port of entry, Supermarket - poultry samples from supermarkets in South Africa, Public data - ST548 genomes uploaded to Enterobase) and presence/absence of antimicrobial resistance (AMR) genes as detected by AMRFinderPlus coloured by AMR class, and presence/absence of plasmid replicons as detected by Abricate.

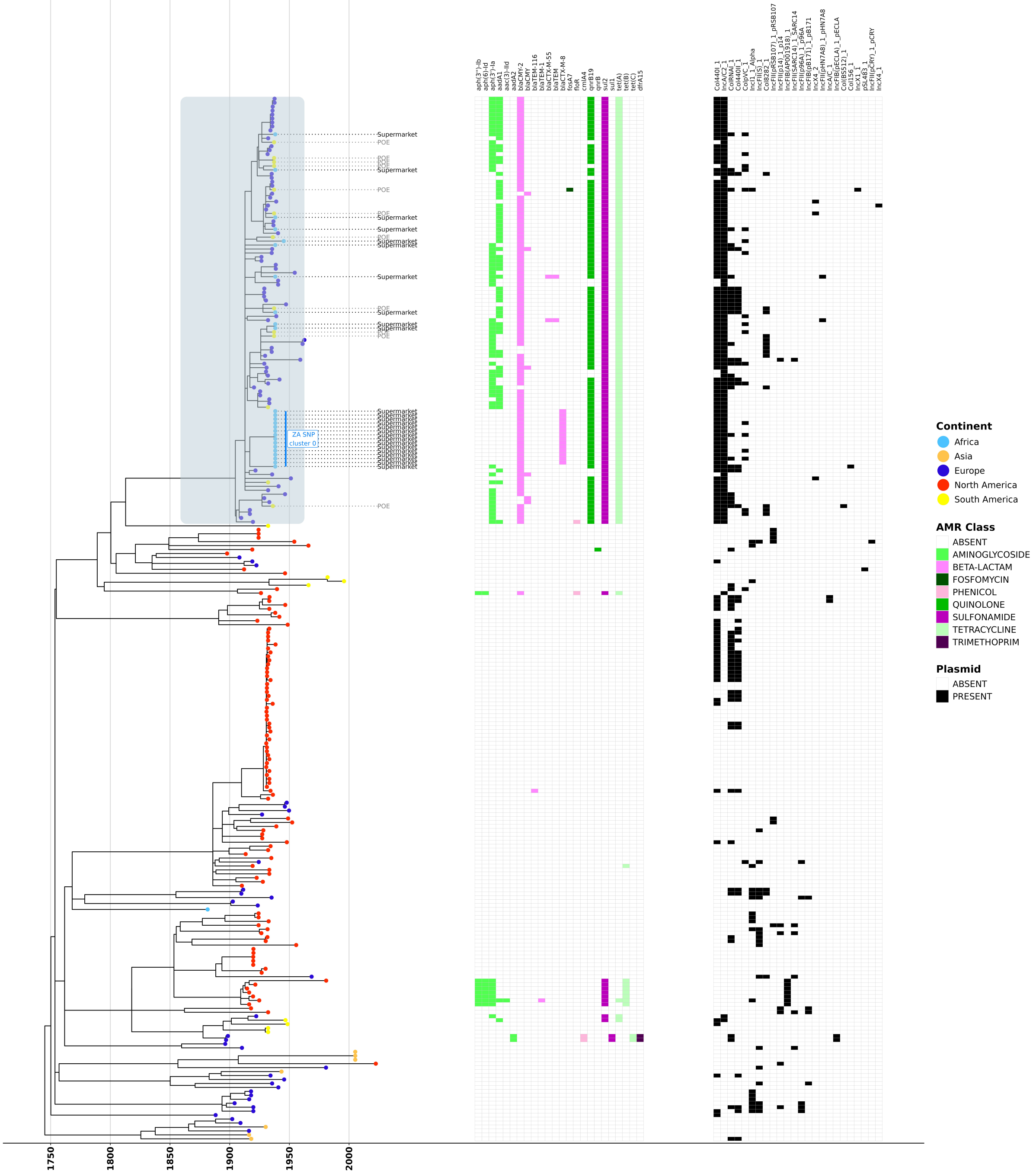


**Fig S4:** Fixed-rate maximum-likelihood (ML) phylogenetic tree constructed using IQ-TREE and RLSD2 comprising all 264 ST548 isolates.Tree tips are coloured by continent of isolation. Heatmap corresponding to tree tips indicate the source of the data (POE - Port of entry, Supermarket - poultry samples from supermarkets in South Africa, Public data - ST548 genomes uploaded to Enterobase) and presence/absence of antimicrobial resistance (AMR) genes as detected by AMRFinderPlus coloured by AMR class, and presence/absence of plasmid replicons as detected by Abricate.

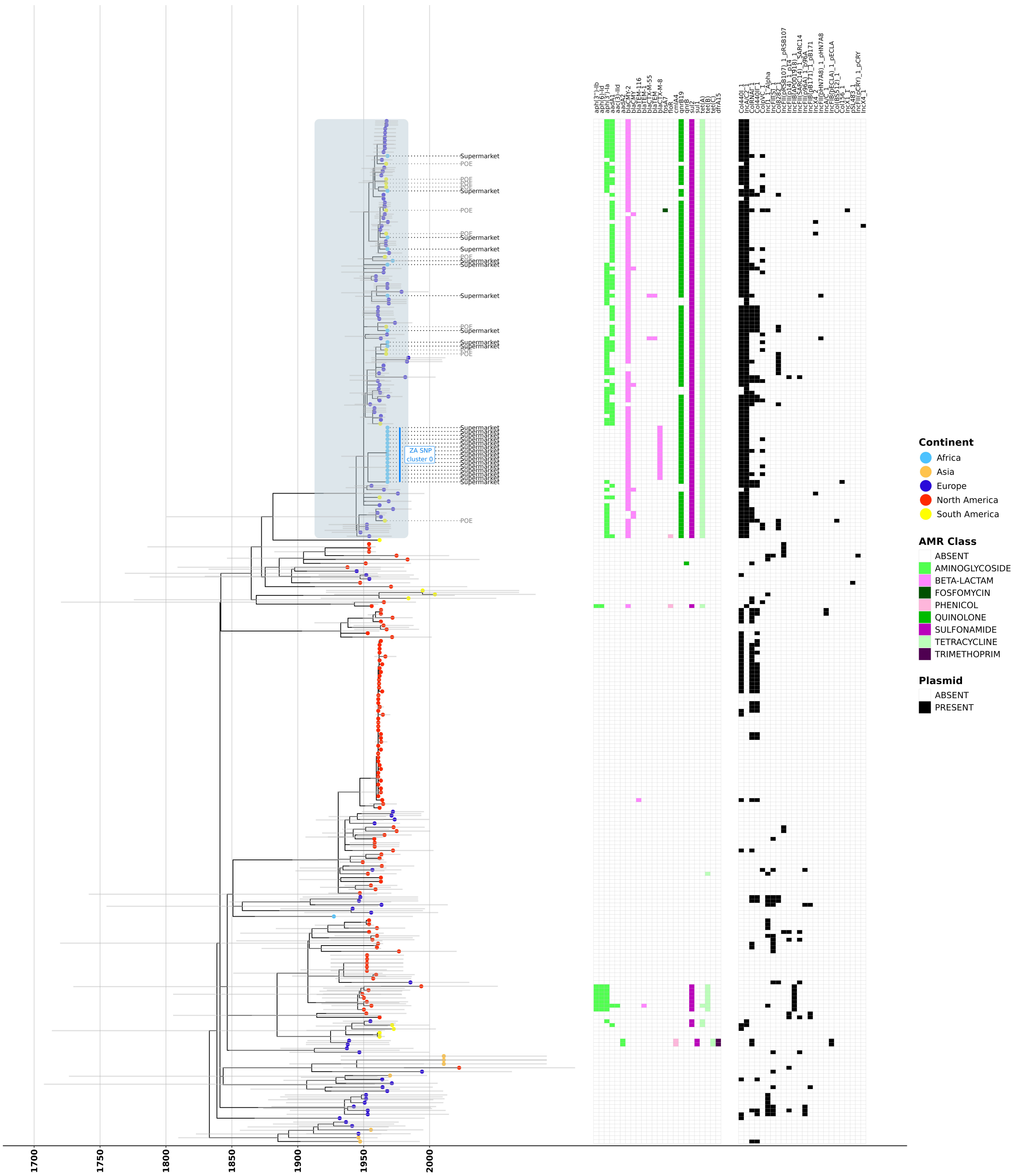


**Fig S5:** LSD2-estimated-rate maximum-likelihood (ML) phylogenetic tree constructed using IQ-TREE and RLSD2 comprising all 264 ST548 isolates.Tree tips are coloured by continent of isolation. Heatmap corresponding to tree tips indicate the source of the data (POE - Port of entry, Supermarket - poultry samples from supermarkets in South Africa, Public data - ST548 genomes uploaded to Enterobase) and presence/absence of antimicrobial resistance (AMR) genes as detected by AMRFinderPlus coloured by AMR class and presence/absence of plasmid replicons as detected by Abricate.

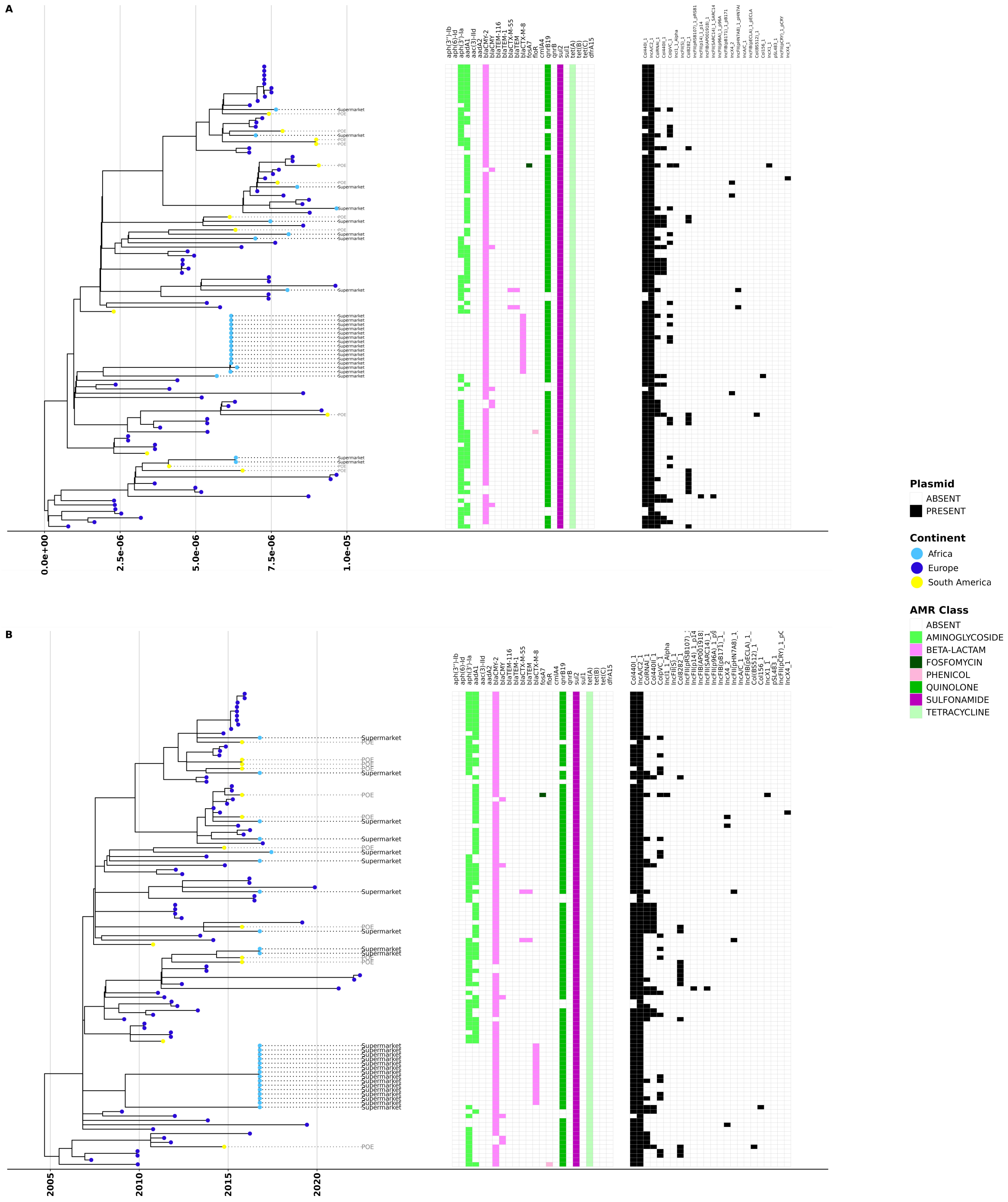


**Fig S6**: Midpoint rooted ML phylogeny (**A**) and LSD2 rate-estimated time scaled phylogeny (**B**) comprising the cluster of 108 high-AMR group isolates (this includes all novel isolates sequenced for this study). Tree tips are coloured by the continent of isolation. Heatmap corresponding to tree tips indicate presence/absence of antimicrobial resistance (AMR) genes as detected by AMRFinderPlus coloured by AMR class, and presence/absence of plasmid replicons as detected by Abricate.

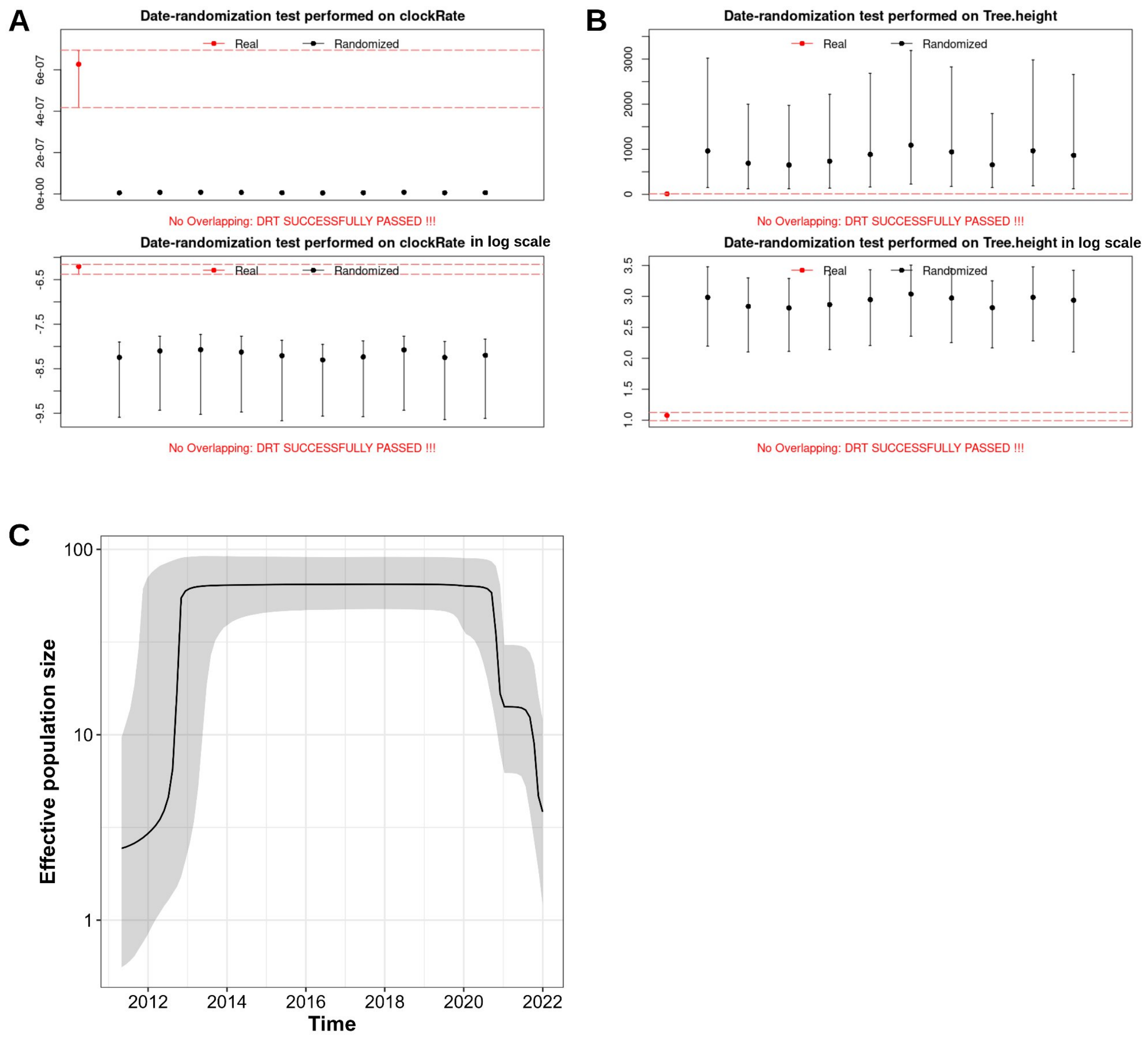


**Fig S7**: Date randomization test performed using \`PlotDRT\` function of R package TipDatingBeast. 10 independent replicates with randomized dates showing no overlap in 95% HPD intervals for clock rate (**A**) and tree height (**B**) when compared to true bayesian phylogenetic reconstruction of 108 high-AMR group isolates of *S.* Minnesota ST548. (**C**) Bayesian skyline plot constructed from Bayesian phylogenetic reconstruction of 108 high-AMR group isolates of *S.* Minnesota ST548. The x-axis shows time in years and the y-axis shows effective population size ($N_e$) . Black line shows median $N_e$ and grey shade represents upper and lower bounds of 95% highest posterior density.

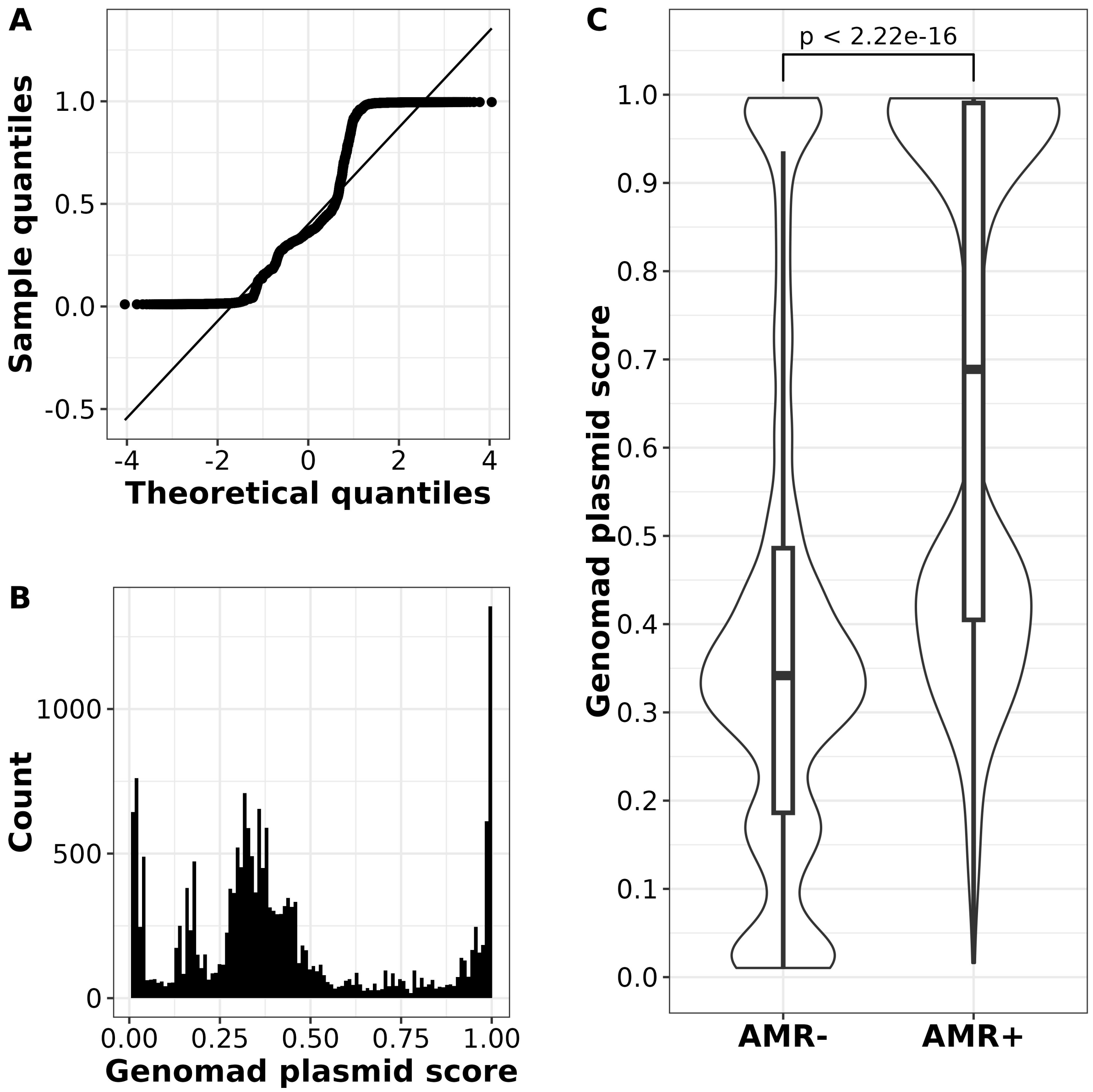


**Fig S8:** Quantile-quantile plot (**A**) and histogram (**B**) of GeNomad predicted plasmid scores of all assembled contigs from 264 ST548 genomes were observed to confirm non-normal distribution of data. Violin plot showing distribution of GeNomad predicted plasmid scores (y axis) for contigs not harbouring any AMR genes according to AMRFinderPlus (**AMR-** on x axis) vs contigs harbouring AMR genes (**AMR+** on x axis). Black horizontal line marks the mean. Kruskal Wallis test was performed comparing mean GeNomad plasmid scores between the two groups (AMR- vs AMR+).

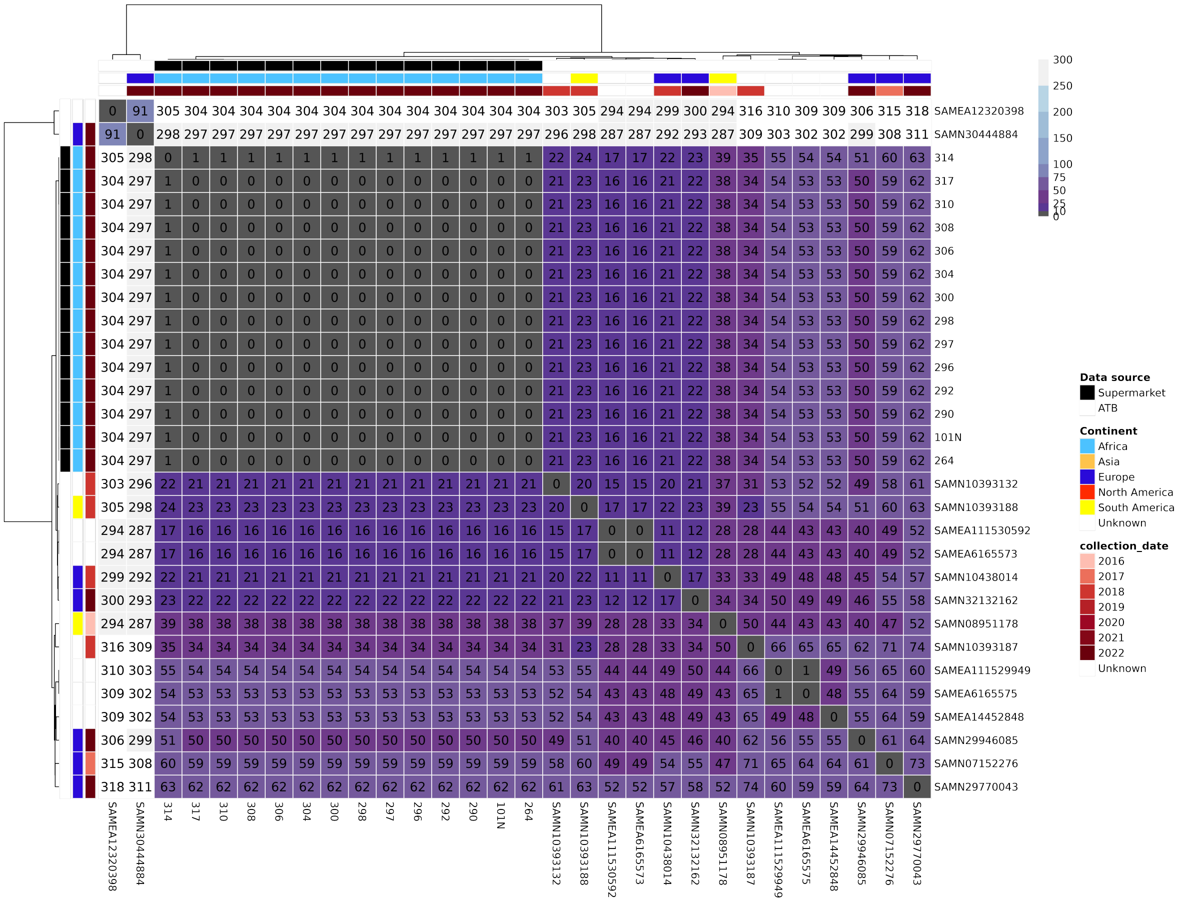


**Fig S9:** All vs all pairwise core SNP distance heatmap (lighter = greater SNP distance) of 14 isolates from ‘ZA SNP cluster 0’ and 16 genomes from AllTheBacteria (ATB). All 30 genomes were positive for blaCTX-M-8 according to AMRFinderPlus. Heatmap is annotated by 1) Source of data (POE - Port of entry, Supermarket - poultry samples from supermarkets in South Africa, Public data - ST548 genomes uploaded to Enterobase); 2) continent of isolation and 3) year of isolation.

### Supplemental table legends

**Table S1**: Internal sample names (`**sample_name**`), metadata (`**Collection year**`, `**Country**`, `**Continent**`, `**Data source**`, `**Isolation source**`), sequence type (`**ST`)** and NCBI accessions (`**biosample_accession**`, `**bioproject_accession**`, `**genome_acc**`) for 36 SEPI genomes + 228 public genomes analysed in this study (n=264).

**Table S2**: Novel isolates sequenced for this study (`**Ref_file**` column) and their corresponding closest publicly available isolate (`**Closest_public_sample**`). The core genome SNP distance between the novel and public isolates are in the `**Distance**` column. `**Collection Year**`,`**Country**`, `**Continent**` and `**Source Type**`  columns show the year, country, continent and source of isolation of the corresponding public isolate.  The biosample, bioproject and genome assembly accessions for the public isolate are in the `**biosample_accession**`,`**bioproject_accession**`,`**genome_acc**` columns respectively.

**Table S3**: Novel isolates sequenced for this study (**Ref_file** column) and their corresponding closest publicly available isolate from South Africa (**Closest_ZA_public_sample**). The core genome SNP distance between the novel and public isolates are in the `**Distance**` column. `**Collection Year**`,`**Country**`, and `**Source Type**`  columns show the year, country (All from South Africa) and source of isolation of the corresponding public isolate. All public ST548 isolates from South Africa were part of bioproject PRJEB39988 (`**Bio Project ID**` column)
